# Relationship of the Microbiome with Neurodegenerative Diseases: Development of AI Tools to Detect Alzheimer’s

**DOI:** 10.64898/2025.12.19.25342688

**Authors:** Alba Pérez-Cuervo, Blanca Lacruz-Pleguezuelos, Diego Coleto-Checa, Laura Judith Marcos-Zambrano, Enrique Carrillo de Santa Pau, Adrián Martín-Segura

## Abstract

This study aimed to develop an artificial intelligence (AI) algorithm capable of distinguishing Alzheimer’s disease (AD) from healthy patients using gut microbiome metagenomics data. To do so, 16S rRNA gene and Whole Genome Shotgun (WGS) datasets available in the literature were utilised. Data was pre-processed and filtered. Then, an initial analysis was done to study classical parameters of microbiome data, such as alpha and beta diversity, as well as to unveil taxonomical differences between the groups that might be influencing disease development, using a unified workflow to enable integration of 16S rRNA gene and WGS data.

Neural Network algorithms were subsequently applied to develop the classifier. First a classical multi-layer perceptron (MLPNN) architecture was tested, which showed limited classification performance, particularly in detecting AD cases. These results were improved using a convolutional neural network (CNN) architecture, due to its better comprehension of hierarchic data like taxa relative abundances sorted following phylogenetic tree structure. In addition, these AI methods were compared with a random forest (RF) classifier, a traditional machine learning algorithm. All models struggled to accurately identify AD cases due to the low number of samples used for algorithm training. Although the RF showed a better performance under such circumstances, observing the evaluation metrics the application of AI to this task reveals promising upon higher amounts of training data. The use of SMOTE, a data augmentation approach, confirmed this assumption improving performance across all models but specifically in the CNN. These results support the utility of microbiome-based diagnostics in AD, but highlight the need for larger, diverse datasets and multi-omics approaches to improve model reliability and uncover disease mechanisms.

## Introduction

### 1. Alzheimer’s disease

Neurodegenerative diseases pose a substantial and growing challenge for global healthcare systems. These diseases, which are typically chronic and long-lasting, affect millions of individuals and require prolonged medical care. Among these conditions, Alzheimer’s disease (AD) is the most prevalent, accounting for 60% to 70% of dementia cases worldwide (1). Ageing is the most significant risk factor for AD, with incidence rates rising exponentially in individuals over 65 years old (2). As life expectancy increases, the number of AD cases is expected to rise dramatically, with over 50 million people currently affected worldwide, a figure projected to triple by 2050. This growing prevalence underscores the urgent need for advances in prevention, treatment, and long-term care strategies (2).

AD is characterised by the progressive deterioration of cognitive functions, especially recent memory, and leads to a gradual loss of functional independence. At a neuropathological level, it is marked by the accumulation of extracellular β-amyloid plaques and intracellular neurofibrillary tangles formed by Tau protein, resulting in synapse loss and neuronal death (3). In addition to these hallmark features, mounting evidence suggests that chronic neuroinflammation plays a key role in the progression of the disease. Activated microglia and elevated levels of pro-inflammatory cytokines have been observed in affected brain regions, contributing to neuronal damage and further amplifying pathological processes (3).

Currently, AD treatments focus on symptom management and slowing disease progression. Cholinesterase inhibitors (donepezil, galantamine, rivastigmine) and the NMDA receptor antagonist memantine offer modest symptomatic relief. Recent advancements include monoclonal antibodies like Lecanemab and Donanemab, which target amyloid-beta plaques to slow cognitive decline. However, these newer therapies carry risks such as brain swelling and bleeding, and their long-term efficacy remains under investigation (4).

In terms of diagnosis, although clinical criteria remain fundamental, complementary tests, including neuroimaging and biomarkers, are increasingly being used to improve the precision of early detection (5). Early diagnosis is critical, as it opens the door to potential preventive strategies before significant cognitive decline occurs. Research suggests that intervening in the preclinical stages of AD could significantly delay or even prevent the onset of symptoms (6).

Despite advancements, achieving reliable early detection remains challenging. Several ongoing studies aim to identify biomarkers and tools that could enhance diagnostic accuracy in the asymptomatic or mild cognitive impairment (MCI) stages of AD. However, progress has been limited, and no definitive approach has yet been established (7). One emerging avenue of investigation is the interaction between the gut microbiota and the central nervous system through the gut–brain axis, which has garnered attention for its potential role in neurodegenerative diseases, including AD.

### 2. Microbiota and Metagenomics

The human microbiota encompasses a vast array of microorganisms residing in various body sites, including the skin, oral cavity, respiratory tract, and gastrointestinal (GI) tract. These microbial communities play essential roles in maintaining physiological processes, such as digestion, immune modulation, and protection against pathogens (8). The gastrointestinal tract harbours a highly diverse microbial community, estimated to include approximately 2000 bacterial species (9). These microbes collectively possess a genetic repertoire that vastly exceeds the human genome, endowing them with the ability to produce metabolites and bioactive compounds essential for host physiology. From early development, this ecosystem shapes immune function, regulates nutrient absorption, and maintains intestinal barrier integrity, highlighting its systemic influence beyond the GI tract (9).

Thus, disturbances in microbiota composition and function, referred to as dysbiosis, have significant consequences for human health. Dysbiosis is characterised by an imbalance in microbial populations, often involving a reduction in beneficial bacteria and an overrepresentation of pathogenic or pro-inflammatory microbes. This state has been linked to various chronic conditions, including metabolic disorders, immune dysregulation, and neurodegenerative diseases (10).

Nonetheless, ageing process significantly influences the composition and diversity of the gut microbiota. As individuals age, there is a notable decline in microbial diversity, accompanied by a reduction in beneficial bacteria such as *Bifidobacterium* and *Lactobacillus*, and an increase in pro-inflammatory microbes like *Enterobacteriaceae* and *Clostridia* (11). These changes are associated with systemic inflammation and a decline in immune function, factors that contribute to the development of age-related diseases, including AD (2).

Understanding the relationship between ageing, dysbiosis, and disease progression has been significantly enhanced by advances in metagenomics. Metagenomics is the discipline that studies the genetic material recovered directly from environmental samples, which in the context of the human body allows for the analysis of entire microbial communities, such as the gut microbiota, without the need for traditional culturing methods. This revolutionary approach enables the identification of microbial species, their relative abundance, and their functional gene profiles. As a result, metagenomic studies provide crucial insights into how microbiota evolves over time and how its dysregulation may contribute to disease (9).

### 3. Microbiota-gut-brain axis

As previously mentioned, the gut microbiota is recognized for its complexity and crucial physiological functions. In addition, growing evidence highlight its impact on the central nervous system (CNS) through the microbiota–gut–brain axis, a bidirectional communication network linking the gut and brain via neural, endocrine, and immune pathways. This intricate system allows gut microbes to influence brain function and behaviour, underscoring their importance in neurological health (12). Key mechanisms of this axis include signalling through the *vagus* nerve, the enteric nervous system (ENS), and the immune system, as well as the production of neuroactive metabolites. Notably, gut bacteria can synthesise or regulate neurotransmitters like serotonin, dopamine, and γ-aminobutyric acid (GABA), which play vital roles in mood regulation and cognitive function (9).

#### The Role of Gut Microbes in Modulating Immune Responses and Neuroinflammation: SCFAs and Tregs vs. SFB and Th17 Cells

Gut microbes produce various metabolites ***(Fig. 1A)***, such as short-chain fatty acids (SCFAs), which are essential for maintaining intestinal barrier integrity, modulating systemic inflammation, and influencing neuronal function (13). SCFAs like butyrate can also promote the differentiation of regulatory T cells (Tregs), which produce interleukin-10 (IL-10) to suppress excessive immune activation and prevent neuroinflammation within the CNS, thus contributing to neuroprotection and reducing the risk of neurodegenerative diseases (14). In contrast, certain gut bacteria, such as segmented filamentous bacteria (SFB), ***(Fig. 1A)*** can induce proinflammatory responses by promoting the differentiation of T-helper 17 (Th17) cells, which produce interleukin-17A (IL-17A), a cytokine implicated in driving CNS inflammation and associated with autoimmune neuroinflammatory conditions like multiple sclerosis (14). This suggests that an imbalance in gut microbiota may exacerbate neuroinflammation and contribute to CNS disorders.

**Figure 1.**
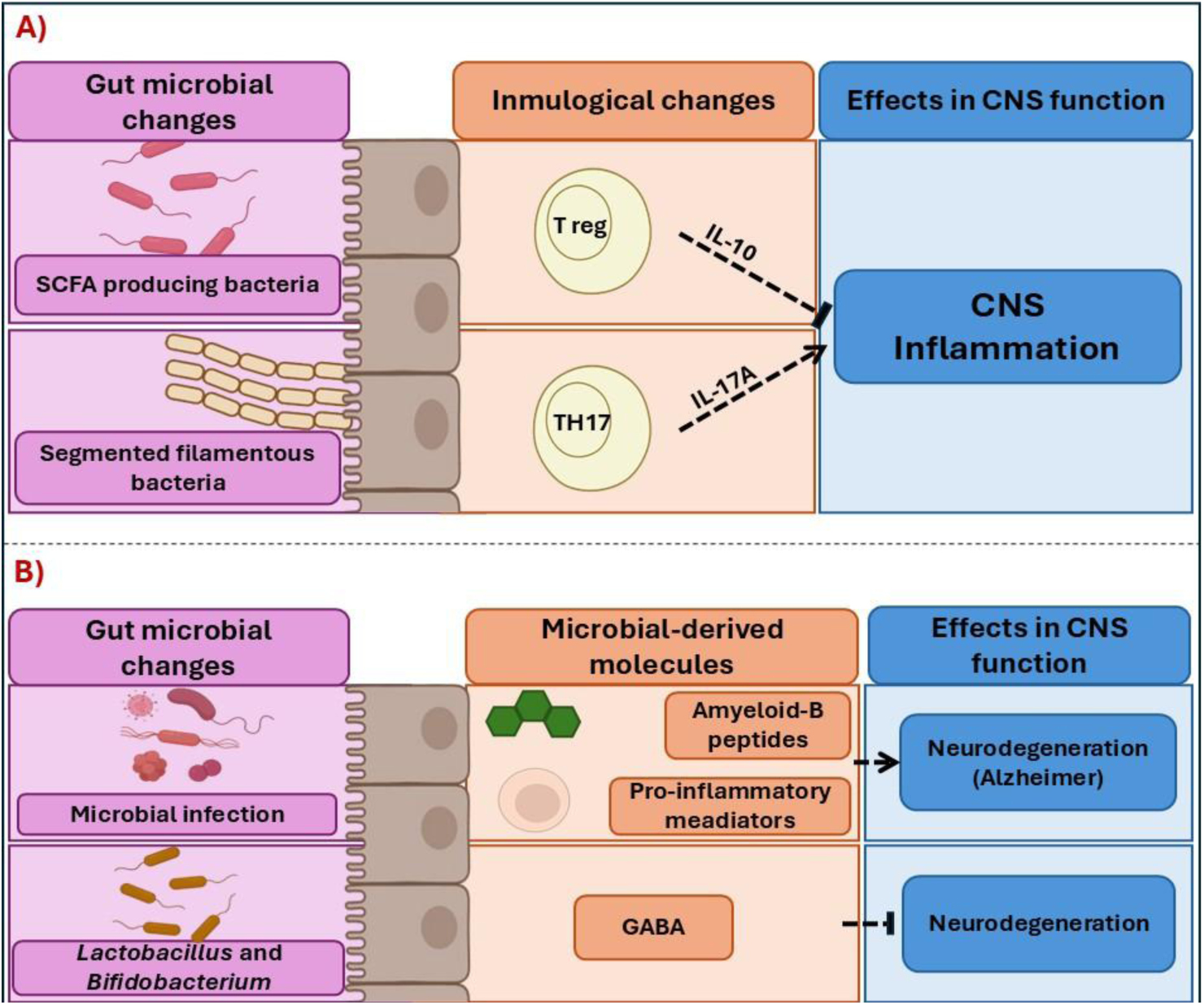
Gut microbiota in fluence on CNS inflammation and neurodegeneration. A) Schematic representation of the gut microbiota’s influence on immune regulation and CNS inflammation. SCFA-producing bacteria promote the differentiation of Tregs, which secrete IL-10, a cytokine associated with anti-inflammatory effects in the CNS. In contrast, segmented filamentous bacteria drive the differentiation of pro-inflammatory TH17 cells, which produce IL-17A, a cytokine linked to CNS inflammation. B) The relationship between gut microbial changes and neurodegeneration through microbiota-derived metabolites. Microbial infections and the presence of beneficial bacteria such as Lactobacillus and Bifidobacterium influence the production of metabolites, including GABA, amyloid-β peptides, and pro-inflammatory mediators. These microbial-derived molecules can impact neurodegeneration and contribute to diseases such as Alzheimer’s disease. Modified from (12) and (14).

#### Amyloid-Beta Peptides, Microbiota, and Neurodegeneration

Moreover, the gut microbiota can influence amyloid-beta (Aβ) aggregation, a hallmark of AD, by modulating host immune responses and altering the metabolic environment (15). Pathogenic bacteria such as *Porphyromonas gingivalis*, a periodontal pathogen, have been implicated in AD progression due to their production of gingipains, proteases that can induce neuroinflammation and contribute to Aβ accumulation (16). Conversely, beneficial bacterial taxa, such as *Bifidobacterium* and *Lactobacillus*, have been associated with neuroprotection due to their role in maintaining gut barrier integrity, reducing systemic inflammation, and producing short-chain fatty acids (SCFAs) that modulate neuroimmune interactions (17). SCFAs, particularly butyrate, have been shown to exert anti-inflammatory effects and enhance microglial function (***Fig. 1B***), potentially mitigating neurodegenerative processes (14).

Additionally, studies in mice indicate that gut microbial dysbiosis and infection can trigger the release of proinflammatory mediators that promote neurodegeneration (14). Elevated systemic inflammation and immune activation, driven by microbial infections, have been linked to increased levels of amyloid peptides in the brain, further accelerating AD pathology. Nonetheless, certain bacterial taxa have been shown to produce amyloid-like proteins that may cross-seed with human Aβ, exacerbating plaque deposition in the brain (15). For instance,

*Escherichia coli* and *Bacillus subtilis* can generate functional amyloids that promote protein aggregation, potentially accelerating neurodegenerative processes (18).

#### GABA and Microbial Influence on the CNS

Another crucial aspect of the gut–brain axis involves the modulation of GABA, the primary inhibitory neurotransmitter in the CNS. Studies have demonstrated that certain gut bacteria, including *Lactobacillus* and *Bifidobacterium*, produce GABA, influencing both the peripheral and central nervous systems (19). GABA plays a pivotal role in reducing neuronal excitability and maintaining the balance between excitatory and inhibitory signalling (***Fig. 1B***). Disruptions in GABAergic signalling have been implicated in anxiety, depression, and neurodegenerative diseases (19). Furthermore, in mouse models, gut-derived metabolites have been shown to influence the function of microglia, the resident immune cells of the brain, which are responsible for synaptic pruning and neuroinflammatory responses. Dysregulation of GABAergic transmission and microglial activity could contribute to cognitive deficits and neuropsychiatric disorders, further emphasizing the role of the gut microbiota in brain function.

#### Gut Microbiota and the hypothalamic–pituitary–adrenal

The hypothalamic-pituitary-adrenal (HPA) axis regulates stress and interacts with the gut microbiome. Early stress can disrupt microbiota and overactivate the HPA axis, leading to long-term effects. Its activation alters gut function and immune responses, contributing to neuroinflammation. In Alzheimer’s, chronic HPA activity and high cortisol promote Aβ and Tau pathology (14).

#### Microbiota in Diagnosis - Current Insights and Challenges

Although gut microbiota has been linked to Alzheimer’s, no diagnostic tool currently uses it clinically. Most findings remain in research. Artificial intelligence (AI) methods like machine learning could help detect microbial patterns in AD, improving early diagnosis and revealing new treatment targets, making this a promising area for future research.

### 4. Artificial Intelligence, Machine Learning, and Deep Learning in Biomedical Research

AI is a broad field of computer science that aims to develop systems capable of performing tasks requiring human-like intelligence, such as pattern recognition, decision-making, and problem-solving (20,21). Within AI, Machine Learning (ML) is a subset that enables computers to learn from data and improve performance over time without explicit programming. Deep Learning (DL), a specialized branch of ML, utilises artificial neural networks (ANNs) with multiple layers to process complex patterns in large datasets (20,21).

In biomedical research, AI and its subfields have become essential for analysing biological data, including the gut microbiome. By employing AI-based models, researchers can uncover correlations between microbial compositions and neurodegenerative diseases such as AD, which may contribute to early diagnosis and personalised treatments (20).

#### Machine Learning Approaches for Microbiome-Based Alzheimer’s Prediction Random Forest: A Non-Neural Network Machine Learning Model

Random Forest (RF) is a machine learning technique commonly applied in biomedical research for pattern recognition and classification tasks. It is an ensemble learning method that constructs multiple decision trees and combines their outputs to improve classification accuracy and reduce overfitting. Each tree in the forest is trained on a random subset of data, and the final prediction is determined through majority voting (for classification) or averaging (for regression). RF is particularly effective in handling high-dimensional biological data, making it a valuable tool for analysing complex relationships between microbiome composition and disease states, including AD (22).

#### Neural Networks and Their Role in Microbiome-Based Alzheimer’s Prediction

ANNs are computational models inspired by the human brain, composed of interconnected nodes (neurons) organized into layers. These networks are particularly effective at capturing non-linear relationships within complex datasets (20,21). One of the fundamental building blocks of ANNs is the perceptron, a simplified model of a biological neuron that processes weighted inputs and generates an output based on an activation function. A perceptron ***(Fig. 2*)** consists of multiple input signals (*X_p1_,X_p2_,…,X_pN_*) that are weighted (*W_j1_,Wj2,…,W_jN_*) and summed. A bias term and a threshold (θ) determine whether the activation function produces an output (*dp_j_*) (23).

**Figure 2.**
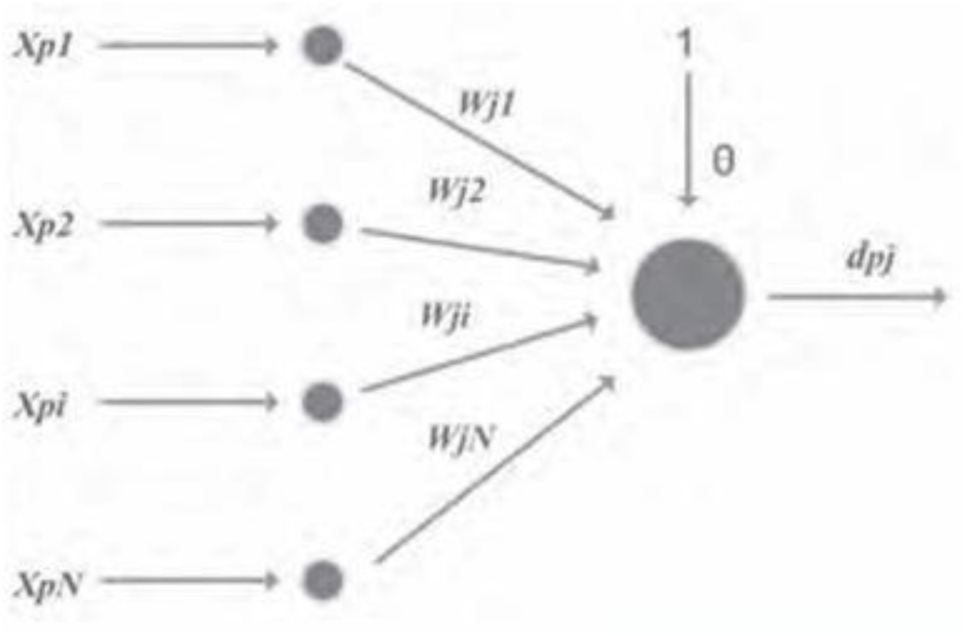
Perceptron architecture. The diagram illustrates the structure of a perceptron, where multiple input signals (*X_p_*_1_,*X_p_*_2_,…,*X_pN_*) are weighted (*W_j_*_1_,*W_j_*_2_,…,*W_jN_*) and summed. A bias term and a threshold (θ) regulate the activation function, determining the final output (*d_pj_*). Modified from (23).

Building upon the fundamental concept of perceptrons, more advanced neural network architectures have been employed to model the relationship between gut microbiome compositions and AD. In particular, Multilayer Perceptrons Neural Networks (MLPNNs), and Convolutional Neural Networks (CNNs) have been utilised due to their ability to capture complex patterns in high-dimensional biological data:

##### i. Multi-Layer Perceptron Neural Networks (MLPNNs)

MLPNNs are feedforward neural networks composed of an input layer, one or more hidden layers, and an output layer, where each neuron is connected to all neurons in the next layer through weighted connections. The hidden layers transform input data using activation functions, introducing non-linearity to capture complex patterns. Learning occurs through backpropagation, which adjusts weights to minimize errors ***(Fig. 3, i***). These networks are particularly useful for microbiome classification, as they can model intricate, non-linear relationships between microbial diversity and disease progression (24).

**Figure 3.**
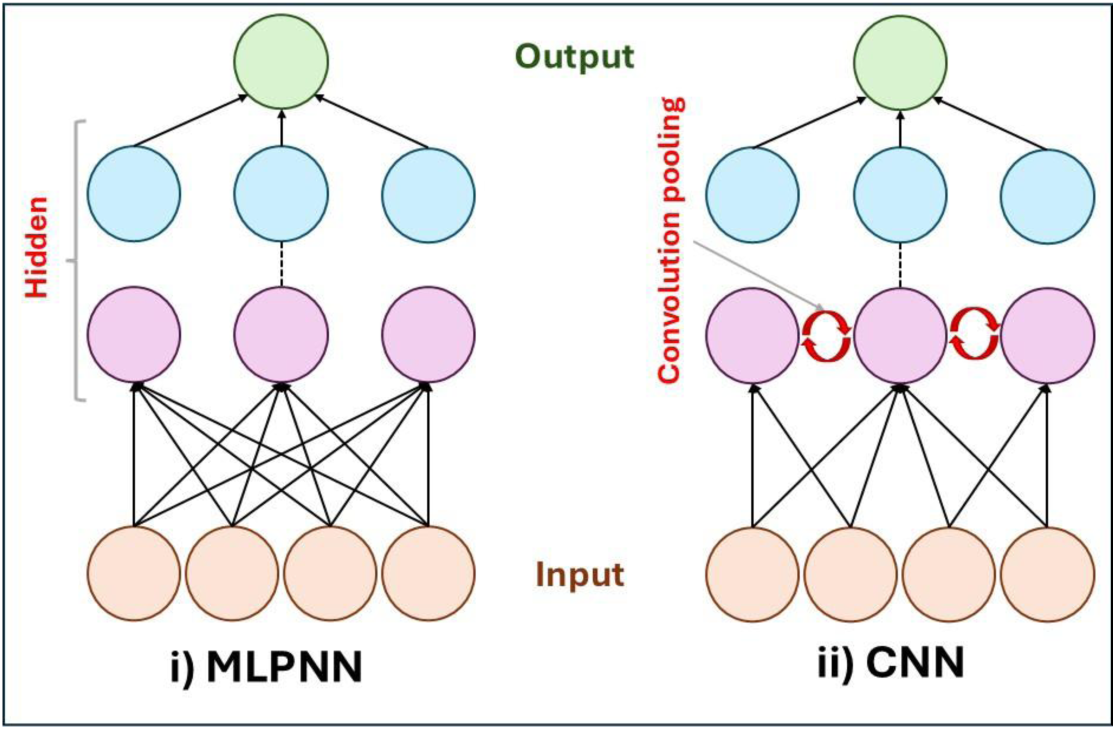
Comparison of neural network architectures used in microbiome-based Alzheimer’s prediction. (**i**) MLPNN is a densely connected neural network with hidden layers that capture complex, nonlinear relationships in microbiome datasets. (ii) CNNs employ convolutional and pooling layers to extract hierarchical features from structured data.

##### ii. Convolutional Neural Networks (CNNs)

CNNs are primarily designed for image recognition but have been adapted for structured and sequential data analysis. They use convolutional layers to extract hierarchical features from input data and pooling layers (e.g., max pooling, average pooling) to reduce dimensionality while retaining key information. The final fully connected layers process these extracted features for classification ***(Fig. 3, ii).*** In microbiome research, CNNs may help identify microbial patterns distinguishing healthy individuals from those affected by AD (25).

By integrating CNNs, MLPNNs, and RF, we can develop AI-driven diagnostic models that analyse gut microbiome profiles to predict AD, facilitating early intervention and personalised treatment approaches.

###### Main objectives

The primary goal of this study was to investigate the relationship between gut microbiome composition and AD by leveraging publicly available sequencing datasets. The study aimed to achieve the following objectives:

1. **Database Compilation and Analysis**: Systematically identify, process, and normalise 16S rRNA and whole-genome shotgun (WGS) sequencing datasets related to gut microbiota in Alzheimer’s disease (AD), creating a curated and quality-controlled database for downstream analyses.
2. **Biomarker Discovery and Therapeutic Insights**: Perform diversity, taxonomic, and statistical analyses to uncover microbial patterns associated with AD, aiming to identify potential biomarkers.
3. **Development of AI-Based Diagnostic Models**: Utilise the compiled microbiome data to develop and train machine learning models (MLPNN, CNN, RF) for the classification of AD versus controls, applying appropriate techniques to address data imbalance and rigorously assessing model performance.

## Materials and methods

### 1. Data Collection

A scoping review was conducted to identify studies investigating the relationship between the gut microbiome and AD. Scientific databases, primarily the National Centre of Biotechnology Information (NCBI) and the European Nucleotide Archive (ENA), were searched with a focus on publicly available 16S rRNA sequencing and shotgun metagenomics datasets. The search strategy utilised a combination of controlled vocabulary terms and free-text keywords related to microbiome composition, neurodegeneration, and sequencing methodologies, along with Boolean operators and filtering criteria (e.g., species specificity, sample type, publication year) to refine the selection (***Fig. 4***). Datasets were included based on the availability of raw sequencing reads in FASTQ format and relevant metadata (e.g., age, sex, cognitive status).

**Figure 4.**
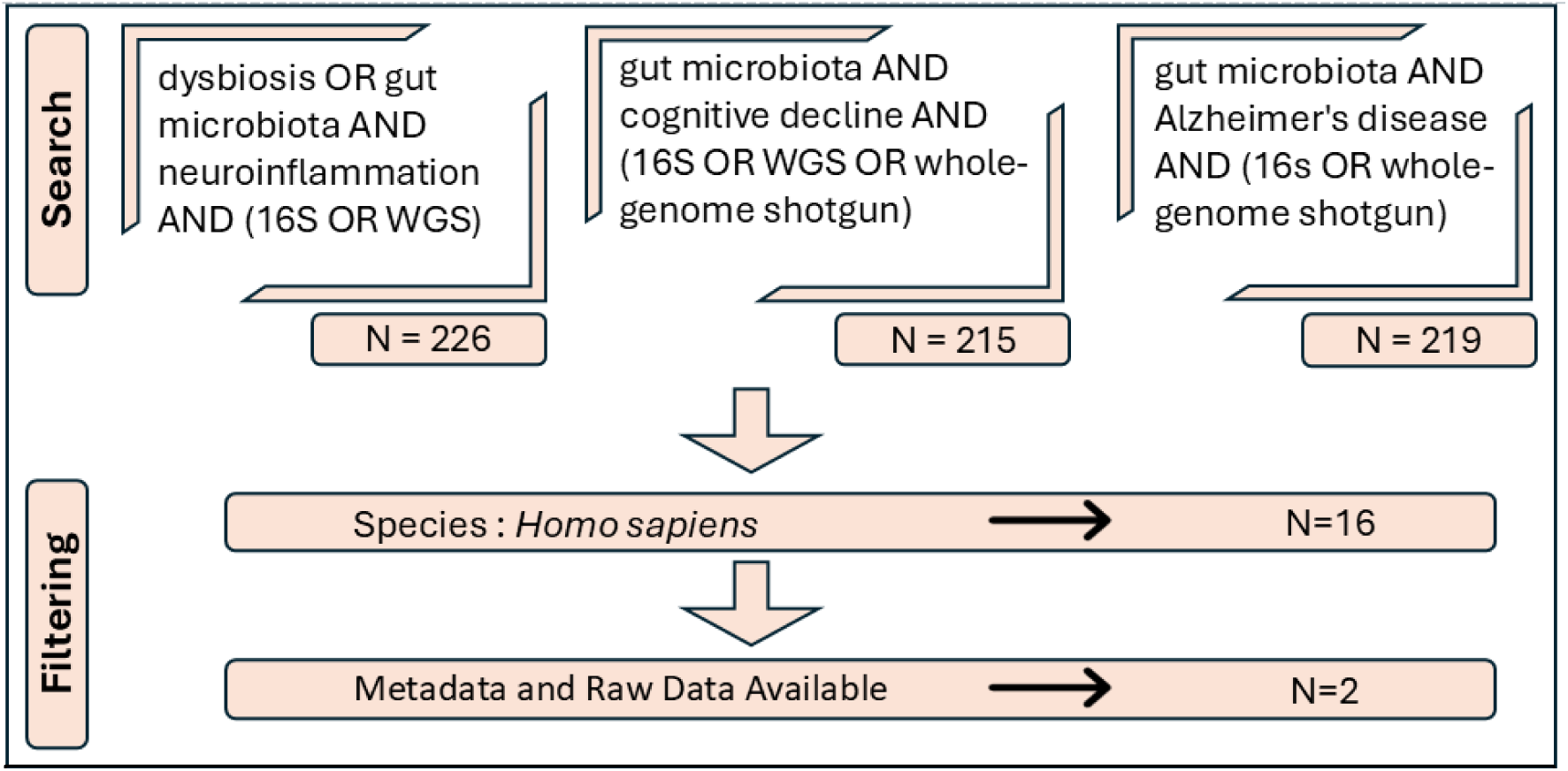
Systematic literature search strategy for studies on gut microbiota and Alzheimer’s disease. The search was conducted using specific query terms related to gut microbiota, neuroinflammation, cognitive decline, and Alzheimer’s disease, incorporating sequencing techniques such as 16S rRNA and whole-genome shotgun (WGS). The initial search retrieved 226, 215, and 219 articles for each respective query. A filtering process was then applied, selecting studies focused on humans, reducing the dataset to 16 studies. Finally, studies providing both metadata and raw data were identified, resulting in a final selection of 2 studies.

### 2. Processing and Quality Control of Sequencing Data

Once the sequencing datasets were selected, the raw sequencing reads were downloaded and processed separately for 16S rRNA sequencing and WGS metagenomics using a custom script (see Data Availability section) tailored for each methodology.

#### 16S rRNA Sequencing Data Processing

For the 16S rRNA sequencing data, preprocessing was performed using QIIME2 (v 2023.2). First, the Conda environment for QIIME2 was created and activated, followed by the installation of the Greengenes2 plugin to enable taxonomic classification. The raw sequencing reads were then retrieved from the selected database and stored in FASTQ format. To facilitate the subsequent analysis in QIIME2, a manifest file was generated using a custom Python script (see Data Availability section), which mapped each sequencing file to its corresponding sample ID and sequencing direction (forward/reverse reads).

The sequences were imported into QIIME2 as paired-end sequences with quality scores, and an initial quality summary report was generated to assess sequencing depth and the overall quality of the reads. To improve data reliability, the DADA2 algorithm was applied for denoising, which removed low-quality reads and filtered out chimeric sequences. This process resulted in the generation of a feature table containing the abundance of amplicon sequence variants (ASVs), along with a set of representative sequences and denoising statistics, which provided insight into the retention of sequences throughout the process.

Once the sequences were cleaned, taxonomic classification was performed using the Greengenes2 database (release 2022.10). The feature table was mapped to the taxonomic reference, generating a classification file that allowed for further taxonomic profiling. The results were visualised through taxa bar plots and additional QIIME2 outputs, such as feature table summaries and sequence tabulations, which were inspected to assess data distribution and metadata integration. All QIIME2 visualisation files were examined using the QIIME2 View platform to verify data quality before proceeding to downstream analyses.

#### Whole-Genome Shotgun Processing

Given that the ultimate goal was to integrate taxonomic classifications from both 16S rRNA sequencing and WGS metagenomics, it was essential to use a common taxonomic reference database ***(Fig. 5)***. Greengenes2 was selected for consistency; however, this database is inherently designed for 16S rRNA-based analyses, and its direct application to WGS data required intermediate processing steps to ensure compatibility.

**Figure 5.**
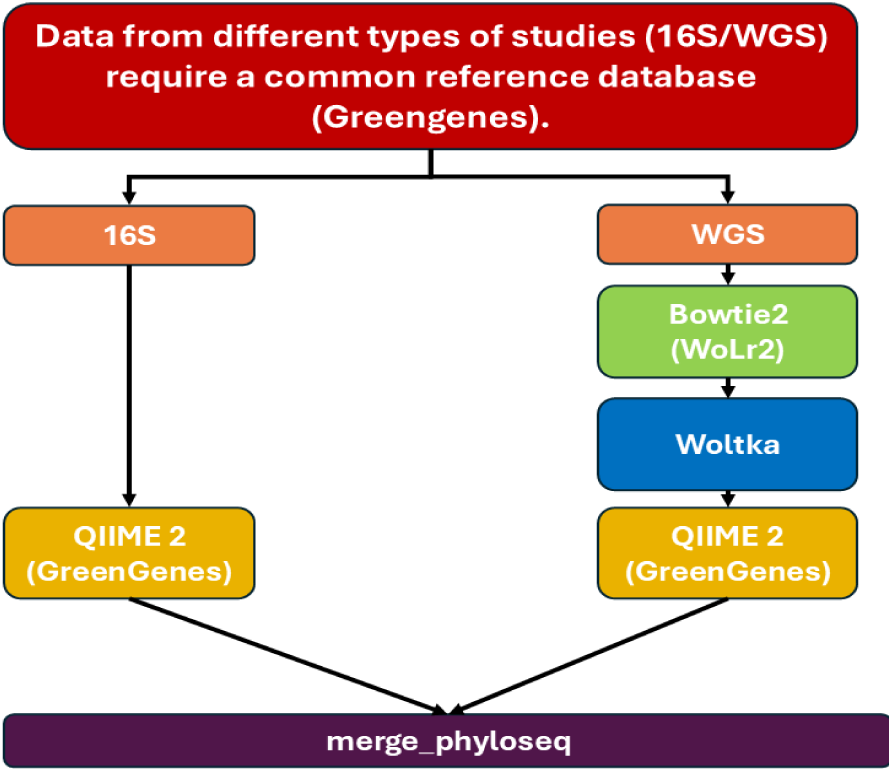
Workflow for integrating 16S and Whole genome shotgun (WGS) sequencing data. A common reference database (Greengenes2) was used for 16S and WGS taxonomic classification to ensure comparability. The figure outlines the preprocessing steps for each methodology, including sequence alignment, taxonomic classification, and integration into a unified dataset using QIIME2.

The initial step in processing the WGS sequencing data involved aligning the raw reads against a comprehensive microbial genome database. For this purpose, the Bowtie2 aligner (v 2.5.3) was employed to map the paired-end sequencing reads to the Web of Life (WoLr2) reference database. A custom Perl script automated the alignment process, ensuring that each sample was correctly mapped while maintaining high sensitivity settings. The output from Bowtie2 consisted of SAM (Sequence Alignment Map) files, each corresponding to an individual sample. These files were then moved to a dedicated directory for downstream processing.

Following alignment, taxonomic classification was performed using Woltka (v 0.1.6), a bioinformatics tool designed for hierarchical genome-based classification. The SAM files were processed within Woltka, generating an Operational Genomic Unit (OGU) table in BIOM format, which represented taxonomic assignments based on whole-genome information. To facilitate organization and subsequent analyses, the OGU files were stored in a structured directory within the computational environment.

To achieve comparability between the WGS and 16S rRNA datasets, a critical filtering step was performed. Since Greengenes2 was originally developed for 16S rRNA gene-based taxonomic assignments, a subset of WGS-derived features that matched Greengenes2 taxa was retained. This process was conducted in QIIME2 (v 2023.2), where the OGU table was first converted into a QIIME2-compatible artefact. Subsequently, only features present in the Greengenes2 taxonomy reference were retained using the Greengenes2 filter-features function in QIIME2. The filtered table was then used for taxonomic classification, aligning the WGS data to the same taxonomic hierarchy as the 16S rRNA sequences.

The final taxonomic assignments for WGS data were stored, which was further formatted into a QIIME2 visualisation file. This step allowed for direct comparison between the taxonomic compositions obtained from 16S rRNA and WGS metagenomics, ensuring consistency across both datasets. The processed taxonomic profiles were subsequently used for downstream comparative and statistical analyses.

### 3. Microbiome Data Processing and Statistical Analysis in R

#### Integration, Normalisation, and Batch Effect Correction of Metagenomic Data

To integrate the taxonomic profiles obtained from 16S rRNA sequencing and WGS metagenomics, both datasets were first converted into phyloseq objects in R(v 4.3.1). The phyloseq package (v 1.44.0) was used to structure the data, incorporating feature tables, taxonomic classifications, and metadata.

The 16S rRNA dataset was processed by importing the feature table and taxonomic assignments using the qiime2R package (v 0.99.6). The taxonomic identifiers were cleaned by removing prefixes to ensure consistency across hierarchical levels. The corresponding sample metadata was then loaded using read_csv from the tidyverse package (v 1.3.2), and column names were standardised to match expected variable formats. Once formatted, the dataset was assembled into a phyloseq object and stored for further analysis.

Similarly, the WGS dataset was processed by loading its feature table and taxonomic classifications via qiime2R. Metadata was incorporated, and an additional variable, *diagnosis*, was created to classify each sample as either *ADs* or *control*. This classification was not determined by our own criteria, but rather followed the diagnostic groupings defined in the original study based on amyloid positivity and other measures such as CSF Aβ42/Aβ40 ratio, centiloid values, and related biomarkers. The dataset was then structured into a phyloseq object.

Before merging both datasets, a study identifier was assigned to each phyloseq object to distinguish the sequencing methodologies. This differentiation was essential for batch effect assessment. The datasets were then combined into a unified phyloseq object, ensuring that taxonomic classifications and sample metadata remained consistent. Since neither dataset contained a phylogenetic tree, a randomized phylogenetic tree was generated using the rtree function from the ape package (v 5.7-1) and incorporated into the merged dataset. Additionally, taxonomic agglomeration was performed at the genus level using the tax_glom function from phyloseq, producing the final dataset for subsequent analyses.

To address potential biases introduced by differences in sequencing depth, rarefaction curves were generated to evaluate sequencing effort across samples ***(Fig. 6).*** A histogram of total read counts per sample was plotted to visualize sequencing depth distribution, and rarefaction curves were constructed using the rarecurve function from the vegan package (v 2.6-4) to examine microbial diversity in relation to sequencing effort. A minimum sequencing depth threshold was set to 40772 reads, corresponding to the sample with the lowest read count. Samples were then rarefied to this uniform sequencing depth using the rarefy_even_depth function from the phyloseq package. This normalisation step ensured that microbial composition comparisons were not biased by sequencing depth disparities. Following normalisation, the presence of batch effects between the datasets was evaluated. Batch effect presence was assessed by transforming the dataset into compositional data using the transform function (microbiome v1.20.0) and performing beta diversity analyses via Multidimensional Scaling (MDS) based on Bray-Curtis dissimilarities. Correction was applied using the adjust_batch function from MMUPHin (v1.8.0), ensuring the preservation of biological signals. The effectiveness of the correction was validated through MDS plots, confirming a reduction in batch effect before proceeding with microbial composition comparisons.

**Figure 6.**
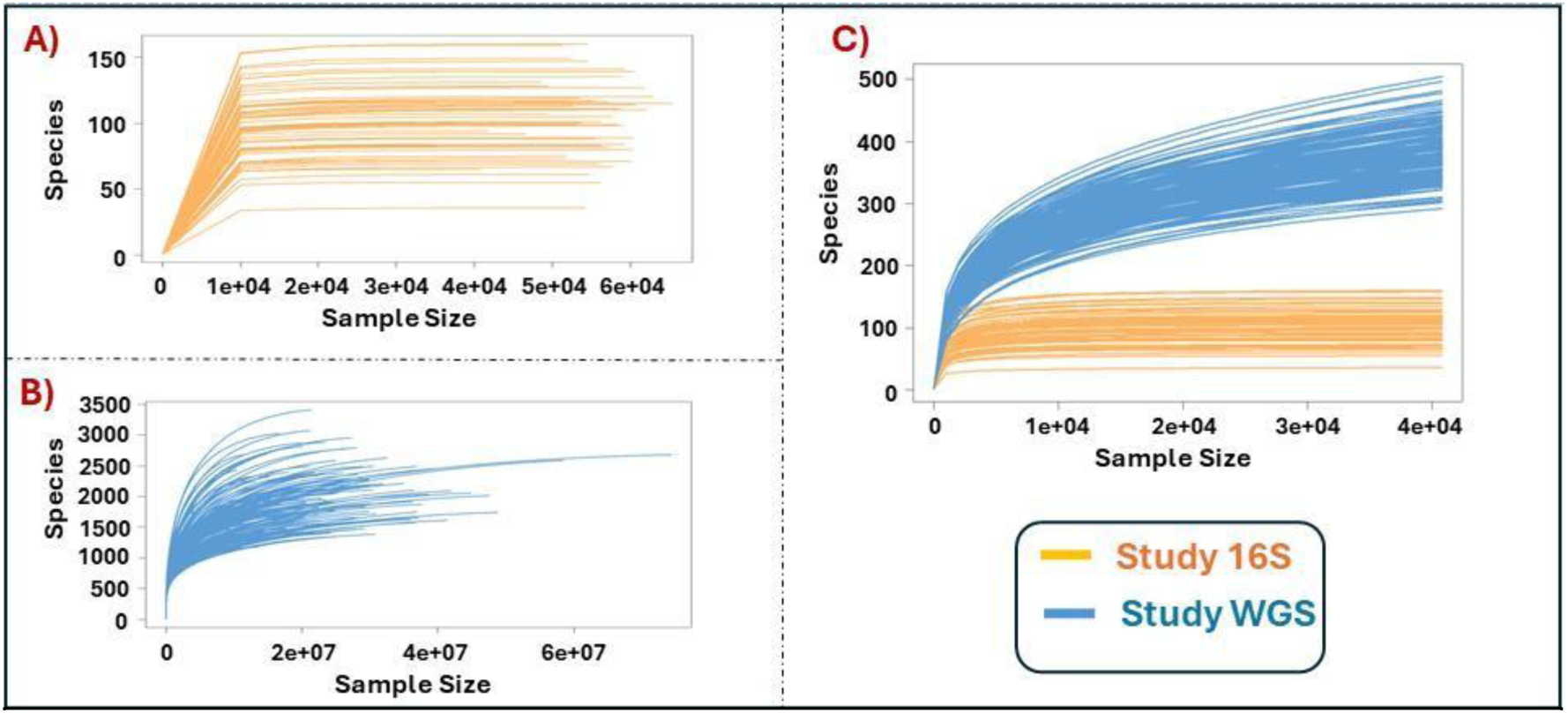
Adjusting different sequencing methodologies to a uniform sequencing depth. (**A**) Rarefaction curves generated from raw sequencing data from the study of 16S, illustrating variability in sequencing depth across samples. (**B**) Rarefaction curves generated from raw sequencing data from the study of WGS, illustrating variability in sequencing depth across samples (**C**) Rarefaction curves after rarefaction to a uniform depth of 40,772 reads per sample, ensuring comparability across samples. The x-axis represents the number of sequences sampled, while the y-axis indicates the number of observed species (richness). This normalisation step mitigates biases due to differences in sequencing depth.

#### Alpha Diversity Analysis

To assess microbial diversity within each sample, alpha diversity indices were calculated from the batch-corrected phyloseq object. The analysis focused on three commonly used alpha diversity metrics: Chao1, Shannon, and Simpson indices. These indices provide complementary perspectives on microbial community structure, capturing species richness and distribution evenness. For visualisation, violin plots were used to illustrate the distribution of alpha diversity values across samples.

#### Taxonomic Filtering and Preprocessing

After computing alpha diversity, taxonomic filtering was applied to remove low-abundance and infrequently observed taxa. The dataset was transformed into compositional data using the microbiome package in R. Taxa with a mean relative abundance below 0.0001 (0.01%) and those present in fewer than 20% of samples were excluded using phyloseq (v1.44.0). Taxonomic names were standardised to correct inconsistencies and ensure uniform classification.

#### Beta Diversity Analysis

Beta diversity was assessed using three distance metrics: Bray-Curtis, Jaccard, and UniFrac, computed from the filtered dataset using phyloseq (v1.44.0). To visualize sample clustering based on microbiome composition, Multidimensional Scaling (MDS) was applied to the computed distance matrices. MDS ordination plots were generated for each distance metric, with samples coloured by diagnostic group and overlaid with 95% confidence ellipses to highlight clustering patterns.

#### Linear Discriminant Analysis Effect Size (LEfSe) for Biomarker Identification

LEfSe was employed to identify differentially abundant microbial taxa between Alzheimer’s and control groups. The analysis involved applying the Kruskal-Wallis test to detect taxa with significant differences across groups (p < 0.05), followed by the Wilcoxon rank-sum test for pairwise comparisons. Linear Discriminant Analysis (LDA) was then performed to assess the effect size of each differentially abundant taxon, using a default LDA threshold of 2.0. The results generated a ranked list of taxa based on their LDA scores, highlighting microbial biomarkers potentially associated with AD.

#### Statistical Analysis

Data normal distribution was assessed using the Shapiro-Wilk test, which determined whether the variables deviated significantly from normality. For variables following a normal distribution, independent two-sample t-tests were conducted to compare patient groups. For variables not following normality, Wilcoxon rank-sum exact tests were used instead. To assess the effects of sex and diagnosis on microbial abundance, a two-way ANOVA was performed, evaluating the main effects and interactions between these factors. To evaluate differences in microbial community compositions between groups, a Permutational Multivariate Analysis of Variance (PERMANOVA) was performed using the vegan (v2.6-4) package in R. PERMANOVA was applied, to Bray-Curtis, Jaccard, and UniFrac distance matrices, with 999 permutations to ensure statistical robustness, enabling the quantification of community dissimilarity across diagnostic groups. Only p-values below 0.05 were considered statistically significant. Exact *p*-values are displayed on the corresponding plots. All statistical analyses were conducted in R (v4.3.1). Figures representing statistical analyses were generated using ggplot2 (v3.4.0) and ggpubr (v0.6.0).

### 4. Machine Learning and Deep Learning Models for Classification

Machine learning, and deep learning model implementation were conducted in Python (v3.11.9). The same preprocessing pipeline was applied to all models designed in this study, ensuring consistency in data handling. The main libraries used included NumPy (v2.0.2) for numerical computations, h5py (v3.11.0) for handling HDF5 files containing microbiome abundance data, and Scikit-learn (v1.5.1) for preprocessing, train-test splitting, classification metrics, and Random Forest implementation. TensorFlow (v2.18.0) and Keras (v3.6.0) were employed for developing deep learning models (MLPNN and CNN), while PyGAD (v3.3.1) was used to implement the Genetic Algorithm for feature selection. Additionally, Imbalanced-learn (v0.12.4) was utilised to perform synthetic data augmentation through SMOTE.

The input files containing microbiome abundance data were processed differently depending on the model used. For MLPNN and RF, the dataset was structured as a two-dimensional matrix where each row corresponded to a patient, and each column represented a specific bacterial taxon. In contrast, CNN required a hierarchically structured format. This structure of the CNN input was generated by arranging taxonomic levels in a way that each level incorporated the cumulative abundance of its preceding level. The first occurrence of each taxonomic category retained the aggregated sum, while subsequent rows were filled with zero values, ensuring a structured representation of microbiome abundance across taxonomic ranks.

The data underwent normalisation using the Centered Log-Ratio (CLR) transformation, a common approach in microbiome analysis to account for compositional data properties. This transformation helps mitigate the impact of differences in sequencing depth across samples, ensuring that relative abundances are properly adjusted for meaningful comparisons. Once the data was normalized and structured, it was split into training and testing sets, with 80% of the data used for training and 20% reserved for testing. Data split was stratified ensuring that both datasets maintained a similar distribution of AD and control cases. Thus, preventing potential biases in model evaluation.

To optimize feature selection and improve classification performance, a Genetic Algorithm (GA) was employed using PyGAD (v3.3.1). GA are evolutionary-based optimisation techniques that iteratively refine solutions by mimicking the principles of natural selection. In this case, the GA aimed to identify the most relevant bacterial taxa contributing to AD classification.

To further enhance model performance, a hyperparameter optimisation process was conducted using GridSearch. This method systematically explored different combinations of hyperparameters to identify the optimal settings for each model. The selection of the best hyperparameters for CNN, MLPNN, and RF was determined based on accuracy and recall metrics. For CNN, the GridSearch process evaluated different learning rates, kernel sizes, dropout rates, and filter sizes for the convolutional layers. Three different validation strategies were tested: without cross-validation, with a 5-fold cross-validation, and with a 10-fold cross-validation. For MLPNN, the GridSearch focused on tuning the learning rate parameter and batch size, evaluating its impact using the same three validation strategies (no cross-validation, 5-fold, and 10-fold cross-validation). For RF, GridSearch was applied to optimize the number of estimators, maximum tree depth, and the minimum number of samples required to split a node and form a leaf. As with the other models, three different cross-validation strategies were considered to ensure robustness in hyperparameter selection.

#### Model Evaluation and Performance Metrics

To evaluate the models, a classification report was generated for each approach, providing key performance metrics such as accuracy, precision, recall, and F1-score. These metrics allowed for a comprehensive assessment of how well each model distinguished between AD and control cases. Additionally, a confusion matrix was computed to analyse the number of true positives, false positives, true negatives, and false negatives, offering insight into each model’s strengths and weaknesses in classification. For the deep learning models (MLPNN and CNN), an additional evaluation was performed using training history visualisations. Plots were generated to track recall and accuracy over epochs, providing a deeper understanding of how quickly each model stabilized and achieved its optimal performance.

#### Synthetic Data Generation

To generate synthetic datasets, the Synthetic Minority Over-sampling Technique (SMOTE) was applied exclusively to the training set in order to balance the dataset. A gradient analysis was conducted by training models with varying levels of synthetic data and evaluating recall and accuracy. The optimal number of additional samples was determined to be 2,400, ensuring improved classification performance while minimizing noise. Once the SMOTE-generated samples were incorporated, the dataset was again subjected to the same preprocessing pipeline, including a new train-test split, a Genetic Algorithm optimisation, and the training of the MLPNN, CNN and RF models.

### Data Availability

The scripts and datasets used in this study are available at <u>GitHub</u>. In case the embedded link fails, the full URL is: https://github.com/albaperezcuervo/TFM

**Table 1.** Summary of bioinformatics libraries and packages used in the study. This table presents an overview of the bioinformatics libraries and packages utilised in the processing, analysis, and visualisation of microbiome sequencing data. It includes the software name, the version used and a brief description of its purpose.

| Library/Package | Version | Purpose |
| --- | --- | --- |
| Ape | 5.7-1 | Generates and manipulates phylogenetic trees in R |
| Bowtie2 | 2.5.3 | Aligns WGS reads to microbial genomes |
| DADA2 | - | Denoises and removes chimeras from 16S sequences |
| Ggplot2 | 3.4.0 | Constructs statistical graphics in R |
| Ggpubr | 0.6.0 | Enhances ggplot2 plots for publication |
| Ggtree | 3.8.0 | Visualises and annotates phylogenetic trees |
| Greengenes2 | 2022.10 | Reference database for 16S taxonomic classification |
| H5py | 3.11.0 | Handles HDF5 files for abundance data |
| Imbalanced-learn | 0.12.4 | Applies SMOTE to address class imbalance |
| Keras | 3.6.0 | Defines and trains deep learning models |
| MMUPHin | 1.8.0 | Corrects batch effects in microbiome data |
| Microbiome | 1.20.0 | Transforms compositional microbiome data |
| NumPy | 2.0.2 | Supports numerical computing in Python |
| Perl (script) | - | Automates alignment and preprocessing tasks |
| Phyloseq | 1.42.0 / 1.44.0 | Structures and analyses microbiome datasets |
| PyGAD | 3.3.1 | Performs genetic algorithm-based feature selection |
| QIIME2 | 2023.2 | Processes and classifies microbiome sequencing data |
| Qiime2R | 0.99.6 | Imports QIIME2 data into R |
| R | 4.3.1 | Statistical environment for data analysis and visualisation |
| Scikit-learn | 1.5.1 | Implements machine learning tools and metrics |
| TensorFlow | 2.18.0 | Builds and trains deep learning architectures |
| Tidyverse | 1.3.2 | Provides tools for data wrangling and visualisation in R |
| Vegan | 2.6-4 | Conducts ecological and diversity analyses |
| Woltka | 0.1.6 | Classifies WGS data taxonomically at genome level |

## Results

### 1) Selected Datasets and preliminary analysis

Following the systematic search and selection criteria ***(Fig. 4***), two sequencing datasets were identified for analysis:

- **16S rRNA sequencing dataset**: *“Altered gut microbiota in Chinese patients with Alzheimer’s disease”*, published in *PLOS ONE* (26). This dataset consists of 60 samples, including 30 AD patients and 30 cognitively healthy individuals (***Fig. 7***).
- **Whole-genome shotgun metagenomics dataset**: *“Gut microbiome composition may be an indicator of preclinical Alzheimer’s disease”*, published in *Science Translational Medicine* (27). This dataset consists of 115 samples, including 49 AD patients and 66 cognitively healthy individuals ***(Fig. 7).***

**Figure 7.**
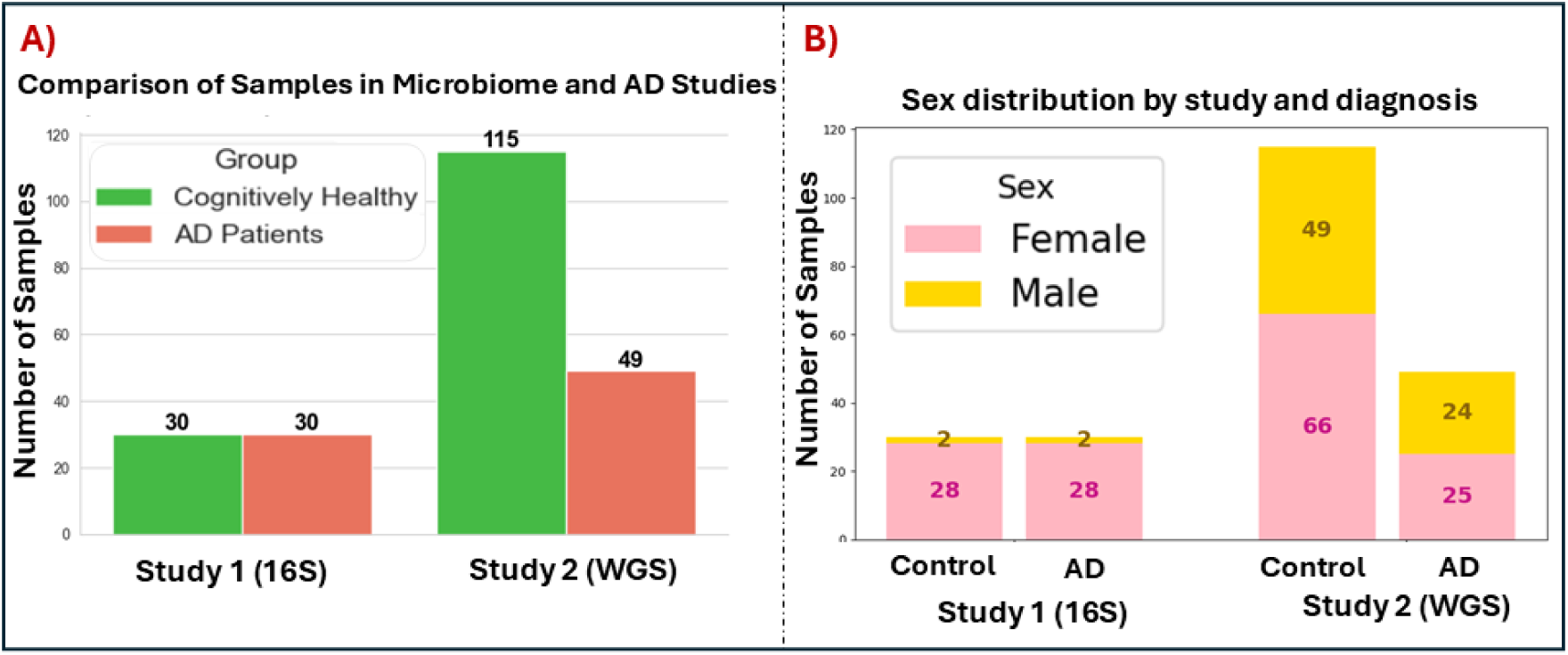
Samples characteristics in microbiome and Alzheimer’s disease studies. (**A**) Comparison of samples distribution between cognitively healthy individuals and AD patients across the two selected studies. (**B**) Sex distribution by study and diagnosis.

An initial assessment was conducted to determine the presence of batch effects resulting from dataset variability. This was achieved by visualising beta diversity via an MDS plot based on UniFrac distances (***Fig. 8A).*** A strong batch effect was evident, with samples clustering primarily according to sequencing methodology rather than biological differences. This distinct separation suggests that technical variations in sequencing introduced a bias, potentially confounding downstream microbial composition analyses.

**Figure 8.**
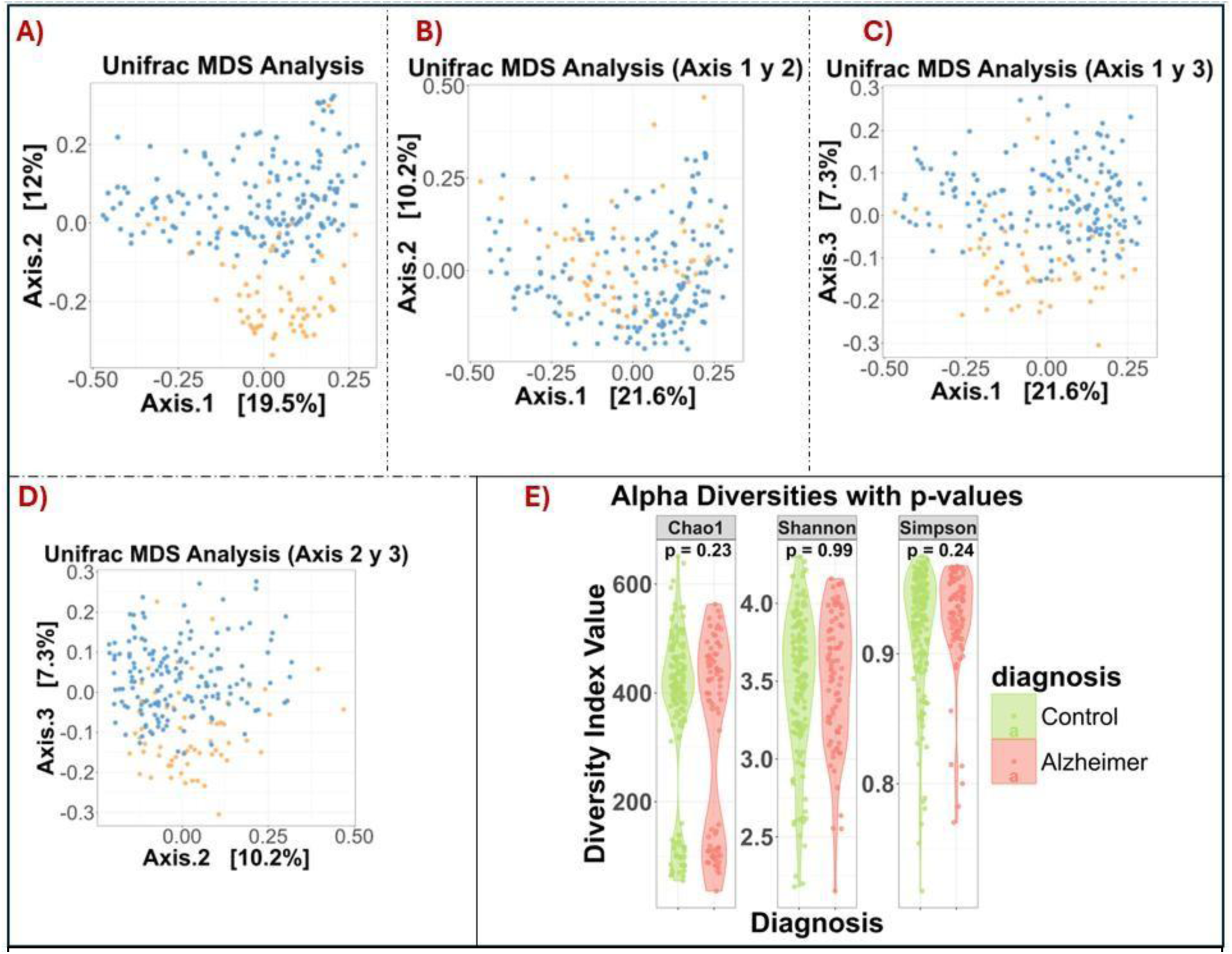
Batch effect correction and alpha diversity comparisons in Alzheimer’s and control Groups. (**A, B, C, D**) Multidimensional Scaling (MDS) plots based on UniFrac distances illustrating the batch effect presence and its correction. 16S or Study 1 samples are depicted in orange and WGS or Study 2 in blue. (A) The initial MDS analysis (Axis 1-2) revealed a strong batch effect. (B, C, D) Graphs showing MDS plots after batch correction (B, Axis 1-2), (C, Axis 2-3), and (D, Axis 1-3). (**E**) Alpha diversity comparisons using Chao1, Shannon, and Simpson indices. Each violin plot represents the diversity distribution within each diagnostic group. P-values were calculated using the Wilcoxon rank-sum test. Exact p-values are shown.

To address sequencing depth disparities, rarefaction curves were used to standardise sequencing effort across samples (see materials and methods section 3), allowing for unbiased comparisons of microbial diversity. However, even after rarefaction, the clustering pattern indicated that batch effects persisted, requiring further correction.

To mitigate this issue, microbial abundances were adjusted using MMUPHin (see materials and methods), ensuring that biological signals were preserved while eliminating technical biases. By examining different axis combinations of de MDS Plots (8B: Axes 1-2, 8C: Axes 1-3, 8D: Axes 2-3), it was determined that MMUPHin mitigated the batch effect, resulting in a more homogenized sample distribution. While a residual batch effect is still observable along Axis 3, it appears reduced compared to the original, uncorrected data. This correction enabled comparisons between Alzheimer’s and control groups, reducing the confounding impact of sequencing methodology.

#### Alpha Diversity Analysis

Next, the analysis focused on alpha diversity, which refers to the within-sample microbial diversity, measuring the richness (the number of distinct taxa) and evenness of species present in a given sample. This metric provides insights into the complexity of microbial communities and helps determine whether diversity differs between conditions, such as AD and healthy controls. To assess alpha diversity, the Chao1, Shannon, and Simpson indices, three of the most used metrics, were employed, revealing varying patterns in microbial richness and evenness between AD and control groups (***Fig. 8E***).

The Chao1 index, which estimates the number of unique species (richness), showed a higher median value in the Alzheimer’s group compared to controls; however, the difference was not statistically significant, suggesting that overall species richness is comparable between conditions. Nonetheless, the Chao1 measure displayed a clear separation into two clusters, likely reflecting residual batch effects, potentially corresponding to samples from different studies. Similarly, the Shannon index, which measures diversity by combining richness and evenness, also exhibited no significant difference between groups, indicating that the overall microbial diversity distribution remains similar across conditions. Finally, the Simpson index, which emphasizes species evenness and dominance, also showed a non-significant difference. Overall, these findings suggest no marked differences in microbial richness and evenness between individuals with Alzheimer’s disease and healthy controls. However, further analyses may be needed to better understand potential variations in community structure.

#### Beta Diversity Analysis

Subsequently, the analysis focused on beta diversity, which measures differences in microbial composition between samples. This metric helps assess whether microbiome profiles significantly vary between diagnostic groups. Unlike alpha diversity, which evaluates species richness and evenness within a single sample, beta diversity examines the overall dissimilarity between microbial communities across different samples. This distinction is essential for understanding how microbial communities vary and for detecting patterns related to specific conditions or groups.

Beta diversity was assessed using Bray-Curtis, Jaccard, and UniFrac distance metrics. Each of these metrics provides unique insights into community dissimilarity. The Bray-Curtis and Jaccard indices focus on shared and unique species between samples, with Bray-Curtis incorporating species abundance and Jaccard concentrating on presence/absence. UniFrac, in contrast, is a phylogenetic metric that evaluates the evolutionary relationships among species, offering a deeper understanding of how microbial communities diverge on a phylogenetic level. By applying multiple distance metrics, we can capture different aspects of microbial community structure, ensuring a thorough analysis of community dissimilarity.

These metrics were visualised through Multidimensional Scaling (MDS) plots, generated for multiple axis combinations (e.g., Axes 1-2, Axes 2-3), to explore potential clustering patterns between Alzheimer’s and control groups. Visually, the MDS plots (***Fig. 9B-D***) do not exhibit a clear separation between Alzheimer’s and control samples. While slight differences in dispersion can be observed, the overlap between groups suggests that, based on these metrics, microbiome composition differences are subtle. The 95% confidence ellipses further highlight this overlap, reinforcing the absence of strong clustering based on diagnosis.

**Figure 9.**
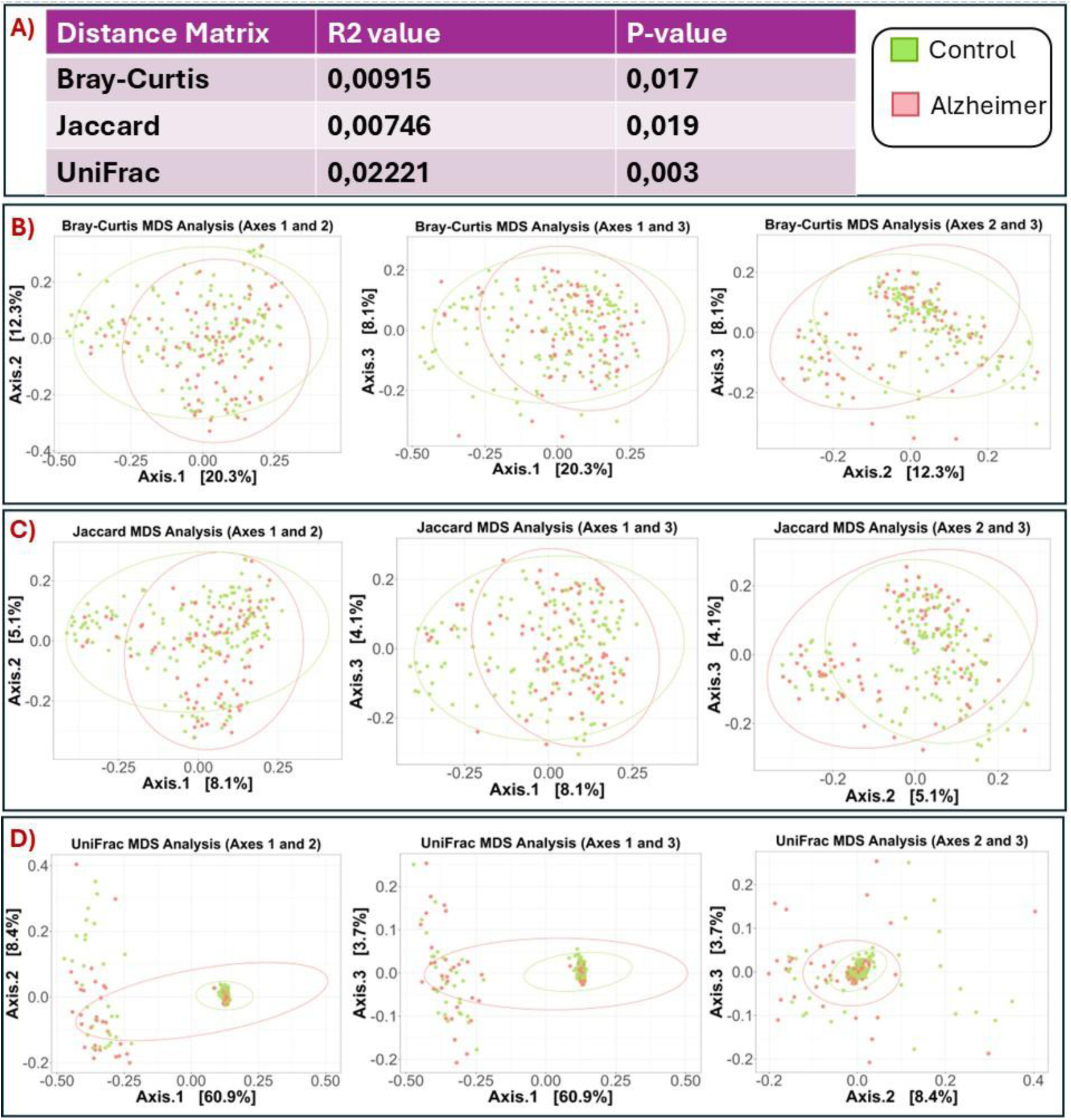
Gut microbiota in fluence on CNS inflammation and neurodegeneration. (**A**) Table summarizing the results of the PERMANOVA test, assessing microbial composition differences between AD and control groups using different distance metrics. (B-D) MDS analysis based on Bray-Curtis (**B**), Jaccard (**C**), and UniFrac (**D**) distance metrics, displaying axes 1-2 (left), 1-3 (centre), and 2-3 (right). Each point represents a sample (green: controls; red: Alzheimer’s). The ellipses indicate 95% confidence intervals.

However, despite the lack of distinct visual separation, statistical analysis using PERMANOVA test revealed significant differences in microbial composition between groups (***Fig. 9A***). The Bray-Curtis distance metric yielded an R² of 0.00915 with a significant p-value (p = 0.017), indicating a slight but statistically significant difference. Similarly, the Jaccard metric showed an R² of 0.00746 with p = 0.019, further suggesting subtle compositional variations. The most pronounced effect was observed with the UniFrac distance, which accounts for phylogenetic relationships, yielding an R² of 0.022221 with p = 0.003, indicating a slightly stronger association between microbial community differences and Alzheimer’s diagnosis. As observed in previous analyses, a residual batch effect appears to persist in the UniFrac MDS plot (***Fig. 9D***), suggesting that some technical variation remains despite the correction applied.

These results suggest that while visual inspection does not reveal strong clustering, statistical analyses detect subtle but significant compositional shifts in microbial communities between Alzheimer’s and control groups. The differences appear to be more pronounced when phylogenetic relationships are considered, as indicated by the UniFrac analysis.

### 2) Microbial Composition and LEfSe-Based Biomarker Discovery in AD

Building on the beta diversity analysis, taxonomic composition was assessed, and LEfSe analysis applied to identify microbial taxa that were differentially abundant in association with AD.

#### Taxonomic Composition

The relative abundance analysis of the top 9 most abundant phyla (***Fig. 10A***) revealed that *Firmicutes* and *Bacteroidota* dominated the microbial communities in both Alzheimer’s and control groups, followed by other phyla such as *Proteobacteria*, *Actinobacteriota*, and *Verrucomicrobiota*. The overall microbial composition appeared similar across groups, with no evident shifts in major phyla. However, statistical testing identified significant differences for specific phyla. The Wilcoxon rank-sum test revealed that *Firmicutes* (p = 3.66 × 10⁻³²), *Bacteroidota* (p = 7.6 × 10⁻^5^), and *Proteobacteria* (p = 2.00 × 10⁻^3^) were significantly different between Alzheimer’s and control groups, suggesting potential taxonomic shifts at higher taxonomic ranks (***Fig. 10B***).

**Figure 10.**
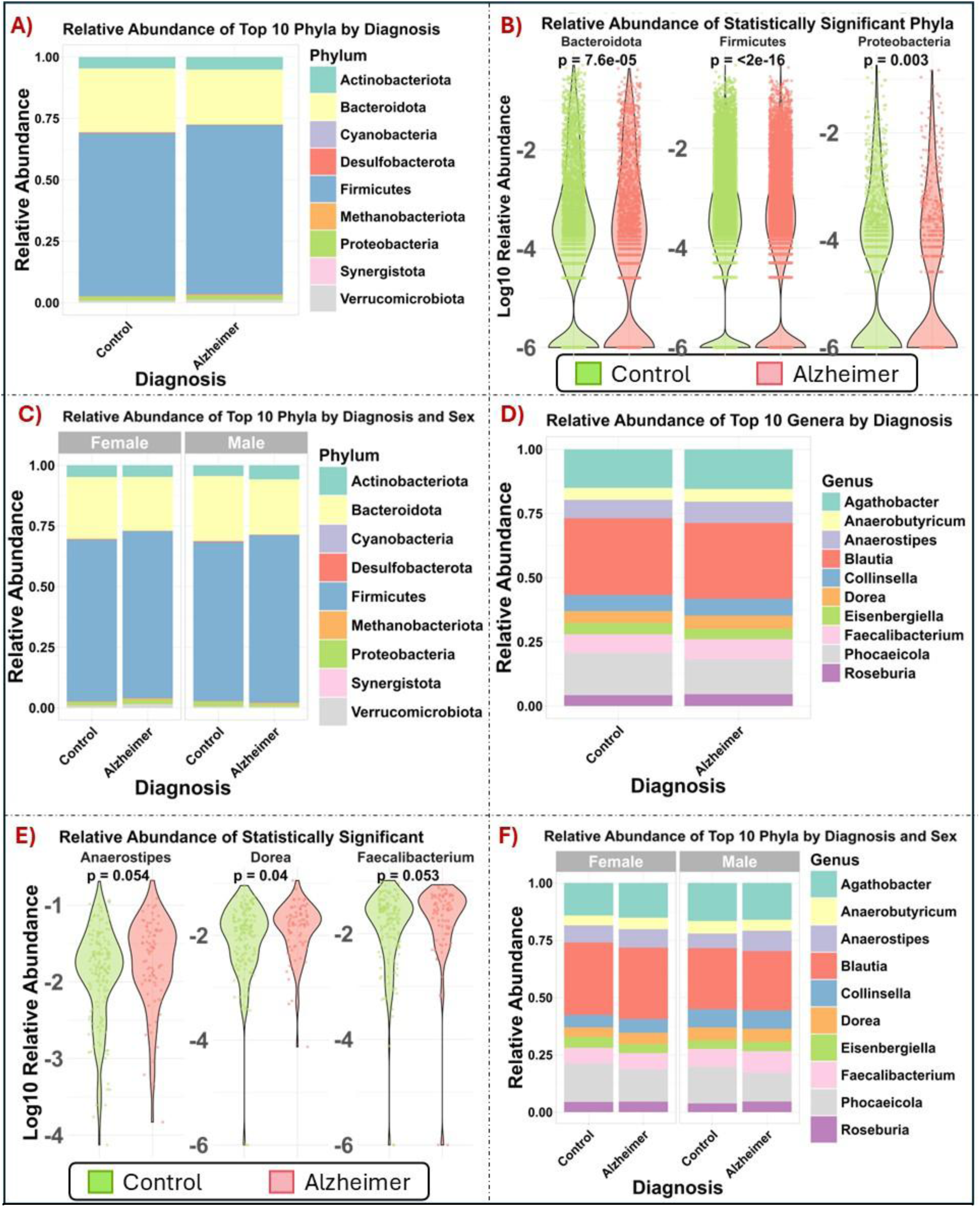
Taxonomic composition analysis. (**A**) Stacked bar plot depicting the relative abundance of the 9 most abundant bacterial phyla in Alzheimer’s and control groups. (**B**) Violin plots representing the phyla that showed significant differences in relative abundance between groups, extracted from the top 9 phyla shown in (A). (**C**) Stacked bar plot showing the relative abundance of the top 9 most abundant bacterial phyla in Alzheimer’s and control groups, stratified by sex (female and male). (**D**), (**E**), and (**F**) correspond to (A), (B), and (C), respectively, displaying the relative abundance of bacterial genera instead of phyla. P-values were calculated using the Wilcoxon rank-sum test. Exact p-values are shown.

These findings remain consistent when considering sex as a variable (***Fig. 10C***), as the relative abundance analysis shows a similar microbial composition across all sex and diagnosis groups, with *Firmicutes* and *Bacteroidota* remaining dominant. A two-way ANOVA test was performed to assess sex influence on microbiota composition. However, the results indicated that neither sex, nor its interaction with diagnosis significantly influence microbial composition at the phylum level.

After analysing the microbial composition at the phylum level, differences at the genus level were examined using the same methodological approach (***Fig. 10D***). Grouping the data at this finer taxonomic resolution, microbial communities in both Alzheimer’s and control groups were found to be dominated by genera such as *Blautia*, *Agathobacter*, *Faecalibacterium*, *Anaerostipes* and *Phocaeicola*.

While the overall composition remained relatively similar, certain genera, such as *Phocaeicola* and *Anaerostipes*, showed changes in relative abundance between groups. Further statistical testing using the Wilcoxon rank-sum test, identified *Dorea* as the only genus showing a statistically significant difference, while *Faecalibacterium* and *Anaerostipes* had p-values close to the threshold for significance, suggesting potential trends worthy of further exploration (***Fig. 10E***). These results indicate that although broad microbial patterns remain largely conserved, specific genera may undergo taxonomic shifts in association with AD.

Finally, sex was considered as an additional factor in the genus-level analysis. The overall microbial composition remained consistent across sex and diagnosis groups, with *Bacteroides*, *Blautia*, *Agathobacter*, *Phocaeicola*, *Faecalibacterium*, *Collinsella*, and *Dorea* as the most abundant genera. While some genera, such as *Eisenbergiella* , *Anaerobutyricum* and *Roseburia*, showed visible differences (***Fig. 10F***). Two-way ANOVA confirmed that neither sex, diagnosis, nor their interaction had a significant effect on microbiota composition; consequently, non-significant p-values are not presented in Figure 10.

In conclusion, the combined findings at both the phylum and genus levels suggest that certain taxonomic shifts occur in AD and that they are not influenced by sex. Further research at finer taxonomic levels may provide deeper insights into these microbial alterations and their potential role in the disease.

#### LEfSe Analysis of Differentially Abundant Taxa

In order to deepen into taxonomic differences, a LEfSe analysis was performed. LEfSe is a bioinformatics tool designed to detect significant differences in microbial composition across biological conditions by integrating statistical significance with biological relevance. Unlike conventional differential abundance tests, LEfSe applies a three-step approach: first, a Kruskal-Wallis test is used to detect taxa with significant differences across groups; second, a Wilcoxon rank-sum test ensures that these differences are consistent between pairwise comparisons; finally, a Linear Discriminant Analysis (LDA) estimates the effect size of each differentially abundant feature. This combination allows LEfSe to rank taxa not only based on their statistical significance but also on their contribution to group separation, making it particularly useful for biomarker discovery in microbiome studies.

The LEfSe analysis identified microbial taxa that significantly differed between Alzheimer’s and control groups, highlighting potential microbial biomarkers associated with the disease. The cladogram representation ***(Fig. 11***) illustrates the phylogenetic distribution of significantly different taxa, with Alzheimer-associated taxa marked in red and control-associated taxa in green. Several taxonomic groups exhibited differential enrichment, like *Desulfovibrionia* (class), *Erysipelotrichaceae* (family), *Clostridium* (genus), and *Ruminococcus* (genus). Conversely, taxa more abundant in the Alzheimer’s group included *Dorea* (genus), *Peptostreptococcaceae* (family) and *Murimonas* (genus). The hierarchical structure of the cladogram emphasizes that these differences occur at multiple taxonomic levels, suggesting potential alterations in microbiome composition linked to disease status.

**Figure 11.**
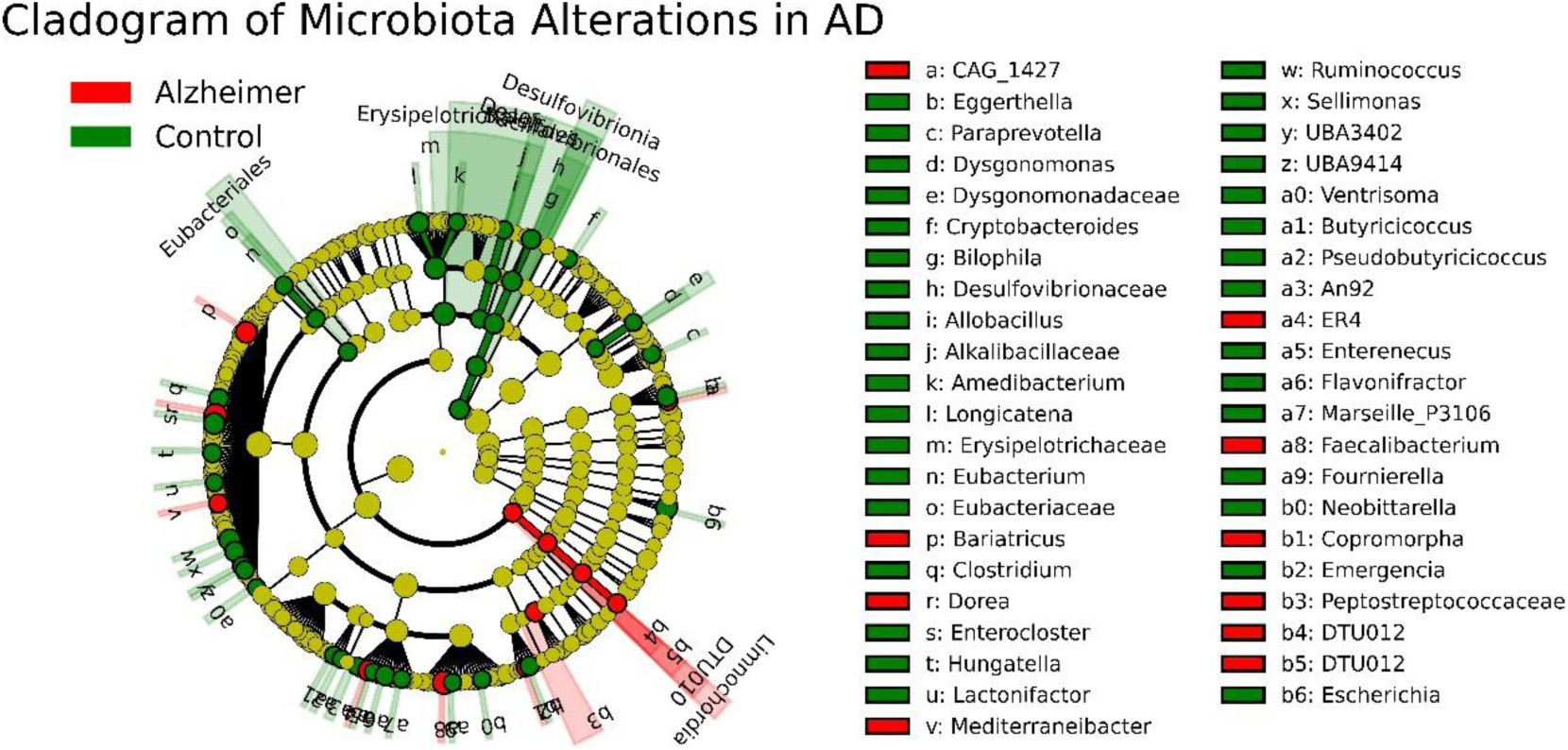
Cladogram of gut microbiota alterations in Alzheimer’s disease Identified by LEfSe. Cladogram representation of taxa with significant differences between Alzheimer’s (red) and control (green) samples. The phylogenetic tree displays hierarchical relationships among taxa, with nodes indicating differential abundance.

The LDA score plot ***(Fig. 12)*** provides a quantitative ranking of differentially abundant genus based on their effect size. Genus with positive LDA scores (green) were more abundant in control samples, while those with negative LDA scores (red) were enriched in Alzheimer’s samples.

**Figure 12.**
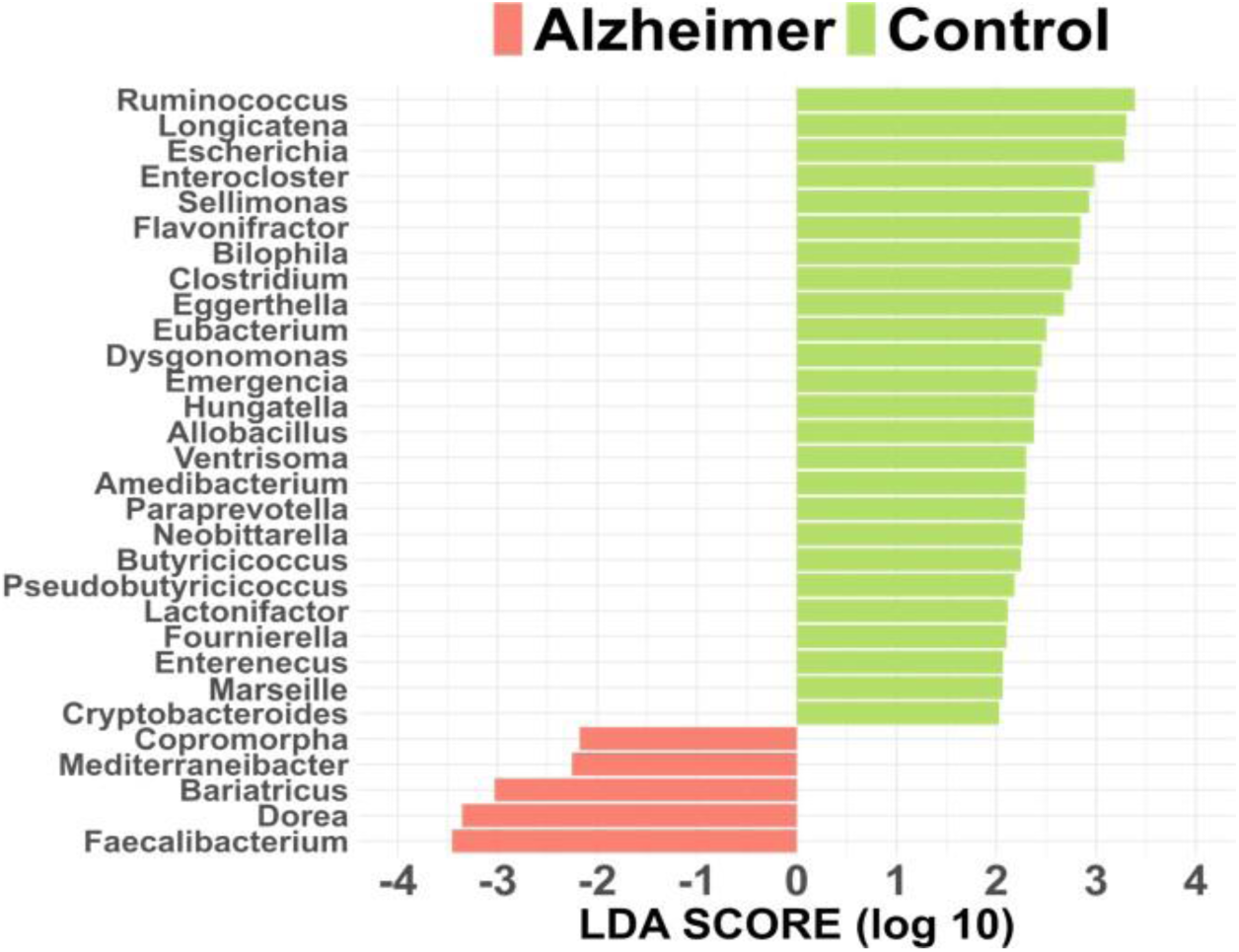
LDA score plot of Differentially Abundant Taxa in Alzheimer’s by LEfSe Analysis. The graphic shows the logarithmic LDA score (log10) of taxa with significant differences between groups. Positive scores (green) represent taxa enriched in control samples, while negative scores (red) indicate taxa enriched in Alzheimer’s samples.

The most discriminative genera enriched in controls included *Ruminococcus, Longicatena, Escherichia, Enterocloster, Sellimonas,* and *Flavonifractor,* while *Faecalibacterium, Dorea,*

*Bariatricus, Mediterraneibacter, Copromorpha,* and *Cryptobacterioides,* were more prominent in Alzheimer’s samples. These findings confirm that shifts in microbial composition are associated with Alzheimer’s pathology, with certain taxa being reduced or enriched depending on disease status.

### 3) Machine Learning and Deep Learning Models for Classification

To evaluate the classification performance of different AI and machine learning models, three architectures were implemented: a MLPNN, a CNN, and a RF classifier. The models were trained on microbiome data normalized using the CLR transformation, and feature selection was performed using a Genetic Algorithm (GA). GA are evolutionary-based optimisation techniques that mimic the principles of natural selection. This approach, along with the data pre-processing, evaluation metrics, and other methodological details, is described in section 4 (Materials and Methods). The dataset was divided into training and testing sets with an 80/20 split, ensuring a stratified distribution of Alzheimer’s disease (AD) and control cases. To assess the effectiveness of each model, confusion matrices, classification reports, and training history were analysed.

The first model designed was a MLPNN consisting of six dense layers with 8, 16, 32, 32, 64, and 1 neuron, respectively. The first two layers used ReLU activation, while the subsequent layers employed tanh activation functions to enhance the network’s ability to capture complex patterns. The final output layer used a sigmoid activation for binary classification. To optimise performance, a grid search (CV=None) was performed over learning rate values and batch size. The best configuration used a learning rate of 0.001 and batch size of 10. The model was then trained using the Adam optimizer and a binary cross-entropy loss function. To prevent overfitting, early stopping was monitored, and training was performed for a limited number of epochs to evaluate rapid convergence.

The MLPNN model achieved 20 true negatives and 6 true positives, while 9 control cases were misclassified as AD (false positives) and 10 AD cases were incorrectly identified as controls (false negatives) (***Fig. 13 A***). The classification report indicates that the model’s performance for control cases was moderate, with a precision of 0.67, recall of 0.69, and an F1-score of 0.68. However, its ability to identify AD cases was considerably weaker, with a precision of 0.40, recall of 0.38, and an F1-score of 0.39 (***Fig. 13 B***). The training history shows that the MLPNN model reached high training recall early in the process, but test recall remained relatively low, suggesting overfitting and limited generalisation capacity. To mitigate overfitting, dropout and regularization techniques were applied, but they were not enough to resolve the issue. (***Fig 13. C***).

**Figure 13.**
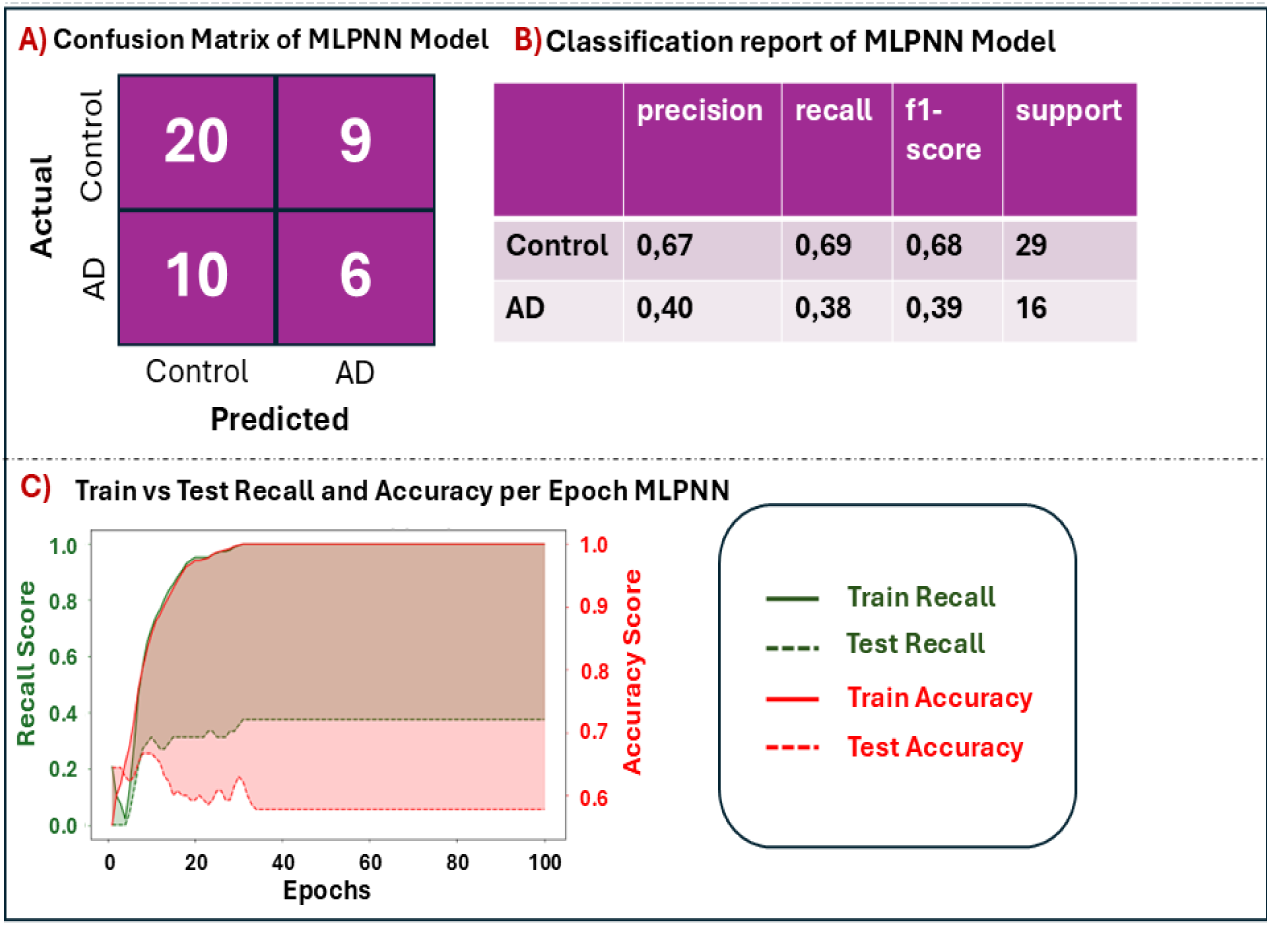
MLPNN model evaluation. (**A**) Confusion matrices llustrating the predictive performance of the MLPNN model before applying data augmentation techniques. (**B**) Classification reports providing precision, recall, f1- score, and support for each class, offering a detailed assessment of the MLPNN model performance. (**C**) Evolution of recall and accuracy metrics throughout the training process, shown as a function of epochs, for the MLPNN model.

The second model was a CNN that included two convolutional layers with 32 and 64 filters, respectively, both using 3×3 kernels with ReLU activation functions. These layers were followed by batch normalisation to stabilize training and max pooling layers to reduce dimensionality. The extracted features were then flattened and passed through a fully connected layer with 128 neurons using a tanh activation function. A dropout layer of 0.5 was added before the final sigmoid activation, which produced the probability of a sample belonging to the AD or control group. To optimise performance, a grid search (CV=None) was performed over key hyperparameters. The final model used a kernel size of 3×3 and a dropout rate of 0.5 and was trained using the Adam optimizer with a learning rate of 0.0001, and a learning rate reduction mechanism was implemented to adjust dynamically based on training loss. Training was conducted for 100 epochs with a batch size of 20, ensuring adequate convergence. ***(Fig. 14)***. In terms of performance, the CNN correctly identified 26 control cases (true negatives) and 6 AD cases (true positives) but misclassified 10 AD cases as controls (false negatives) and 3 control cases as AD (false positives) (***Fig. 14 A***). In terms of precision and recall, the CNN exhibited higher precision for AD cases (0.67) compared to the MLPNN, but a recall of only 0.38, resulting in an F1-score of 0.48. For control cases, the CNN performed better, with a precision of 0.72, recall of 0.90, and an F1-score of 0.80, indicating that while the model was more reliable in identifying controls, it struggled with detecting AD cases (***Fig. 14 B***). The training history shows that the CNN rapidly reached near-perfect training recall, but test recall fluctuated significantly, suggesting potential overfitting. Similar to the MLPNN, dropout and regularization techniques were employed, but these measures did not fully prevent overfitting (***Fig. 14 C***). Compared to the MLPNN, the CNN demonstrated slightly better classification performance for control cases. This may be due to its hierarchical data processing capabilities, the CNN may have aligned well with the hierarchical patterns observed in phylogenetic trees, which could partly explain its improved performance for control classification.

**Figure 14.**
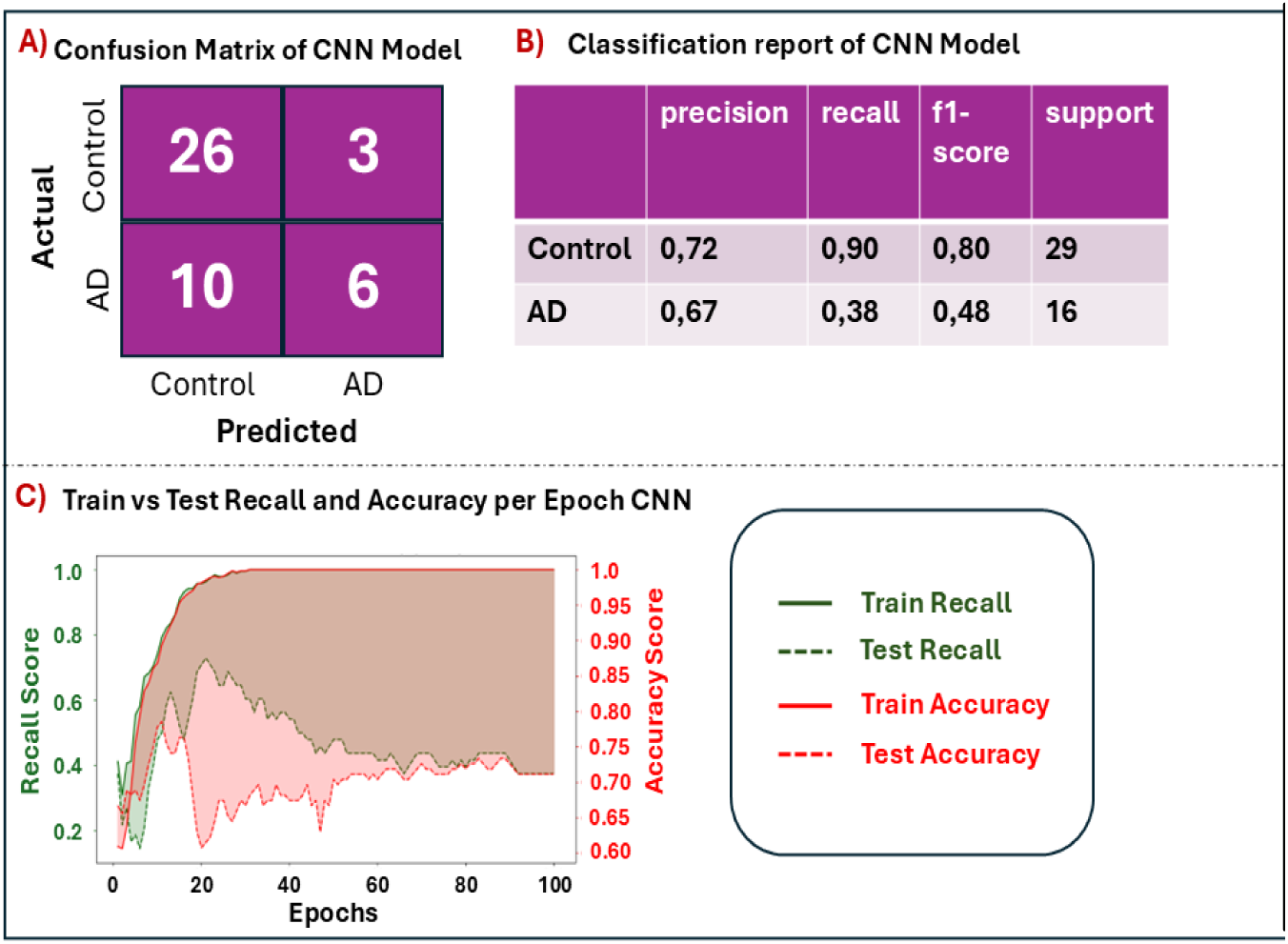
CNN model evaluation. (**A**) Confusion matrices illustrating the predictive performance of the CNN model before applying data augmentation techniques. (**B**) Classification reports providing precision, recall, f1-score, and support for each class, offering a detailed assessment of the MLPmodelperformance. (**C**) Evolution of recall and accuracy metrics throughout the training process, shown as a function of epochs.

The third model was a Random Forest (RF). To optimize performance, a grid search (CV=None) was conducted. The final model used 100 trees, log2 for feature selection, no depth constraint, and a minimum split size of 5. The RF model correctly identified 27 control cases (true negatives) and 6 AD cases (true positives) while misclassifying 10 AD cases as controls (false negatives) and only 2 control cases as AD (false positives) (***Fig. 15 A***). According to the classification report, the RF model had the highest precision for AD cases (0.75), but a recall of only 0.38, resulting in an F1-score of 0.50(***Fig. 15 B***).

**Figure 15.**
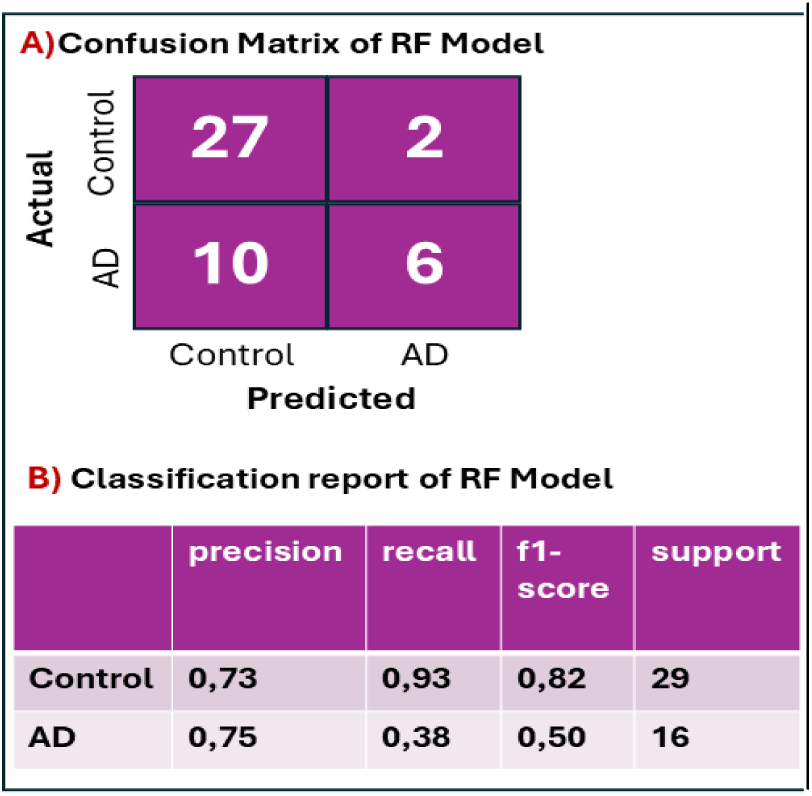
|**Random Forest (RF) model evaluation.** (**A**) Confusion matrices illustrating the predictive performance of the RF model before applying data augmentationtechniques. (**B**) Classification reports providing precision, recall, f1-score, and support metrics for each class, offering a detailed assessment of the RF model performance.

While this represents an improvement over the MLPNN and CNN, the recall remained low, indicating challenges in correctly identifying AD cases. Among the three models, the RF model which served as a baseline comparison against the deep learning approaches, achieved the best classification performance for control cases.

#### Synthetic Data Generation

Given the limited number of samples in the dataset, a data augmentation approach was implemented using the SMOTE, applied exclusively to the training set to enhance model generalisation. SMOTE is a popular technique for handling class imbalances by generating synthetic samples from the minority class. This approach creates new, synthetic instances by interpolating between existing data points, helping the model to generalise better and avoid overfitting. SMOTE was chosen over other methods due to its ability to generate more diverse, realistic synthetic data, which mitigates the risk of overfitting that might arise from simply duplicating minority class instances. This augmentation was crucial for ensuring that the models could achieve better performance on unseen data, enhancing their robustness and generalisation capabilities.

A gradient of simulated data was generated, progressively increasing the total number of synthetic samples up to 4,500, and each model (MLPNN, CNN and RF) was evaluated at different levels of data augmentation to assess its impact on classification performance ***(Fig. 16*).**

**Figure 16.**
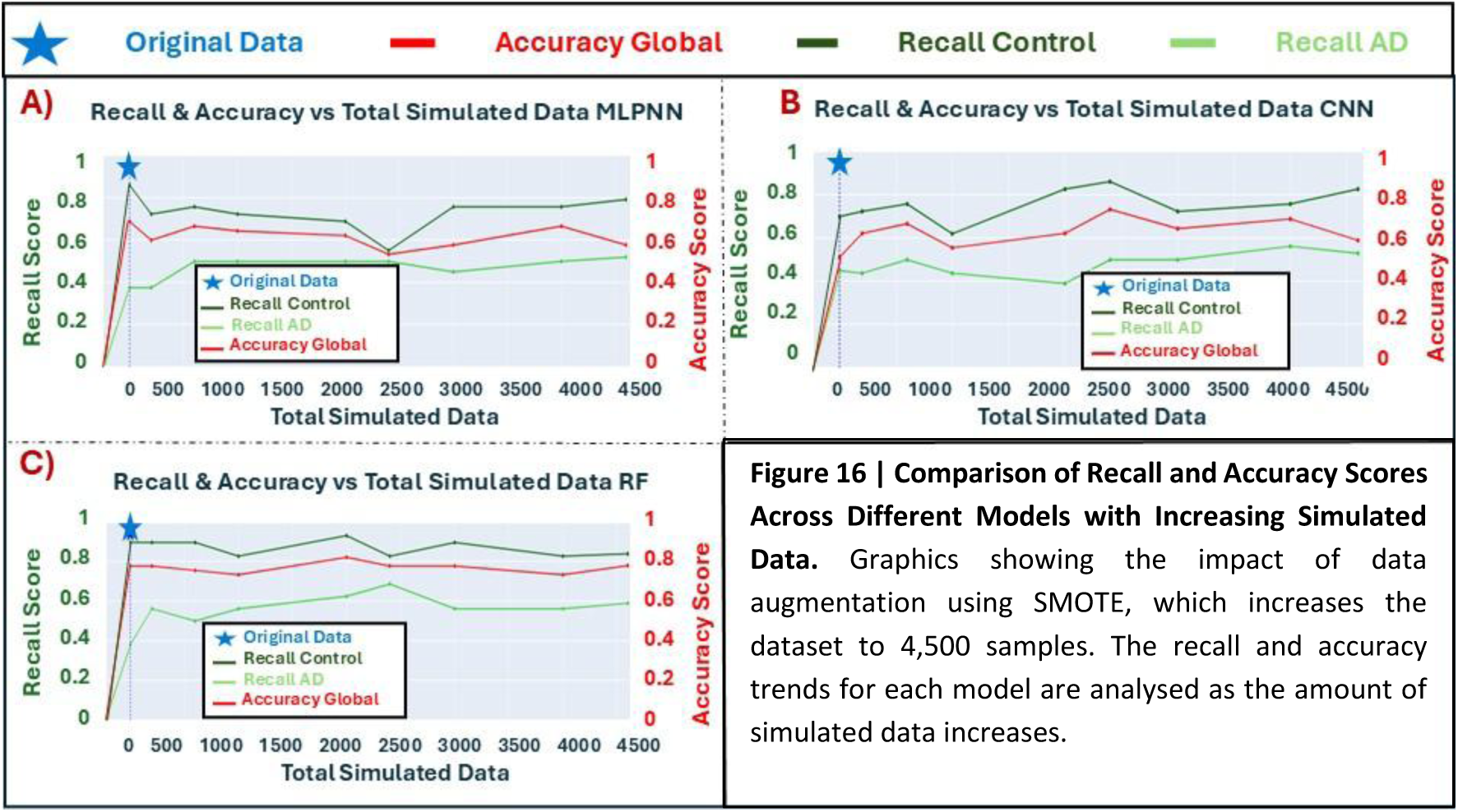
Comparison of Recall and Accuracy Scores Across Different Models with Increasing Simulated Data. Graphics showing the impact of data augmentation using SMOTE, which increases the dataset to 4,500 samples. The recall and accuracy trends for each model are analysed as the amount of simulated data increases.

The evaluation focused on monitoring global accuracy as well as class-specific recall scores for both the control and Alzheimer’s groups. Based on the results, the number of synthetic samples was set to 2,400 (1,200 per class), as beyond this point the performance metrics, particularly for the RF and MLPNN models, remained relatively stable, suggesting limited additional benefit. Furthermore, this level of augmentation corresponded to the point at which the CNN model achieved its highest values across all performance metrics, supporting the choice of 2,400 as a well-balanced augmentation threshold that enhances generalisation without introducing excessive noise.

The classification performance of the models after applying synthetic data augmentation is summarized in ***Fig. 17***. The confusion matrices (***Fig. 18A***) illustrate the classification results for each model. Among the models, the MLPNN model exhibited slightly lower accuracy, correctly classifying 23 control cases and 9 AD cases, with 6 false positives and 7 false negatives. The CNN model showed the best overall performance, identifying 28 control cases and 8 AD cases, with only 1 false positive and 8 false negatives. In comparison, the RF model showed achieved slightly lower performance, correctly identifying 26 true negatives and 7 true positives, but with 3 false positives and 9 false negatives.

**Figure 17.**
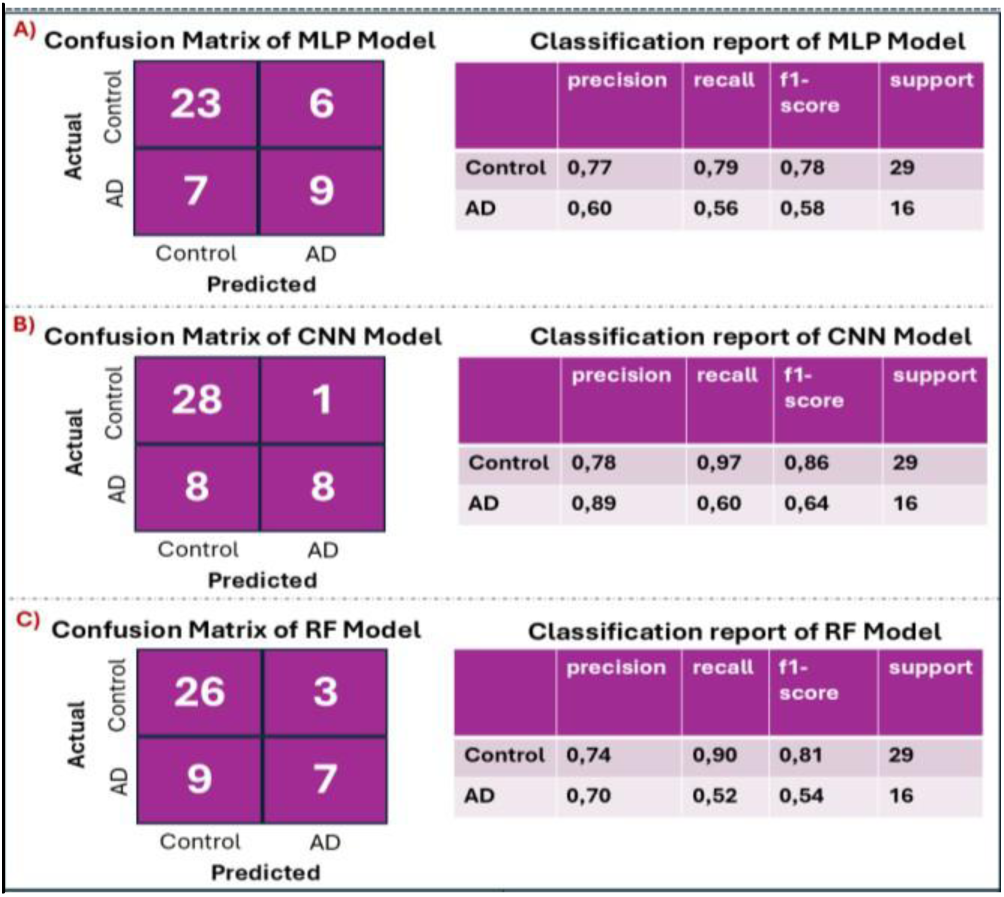
MLP, CNN and RF models’ evaluation after data augmentation. (**A, B, C**) Confusion matrices (left) obtained after augmenting the dataset with an additional 1,200 samples and updated classification reports (right) following data augmentation, reflecting enhancements in model precision and recall.

**Figure 18.**
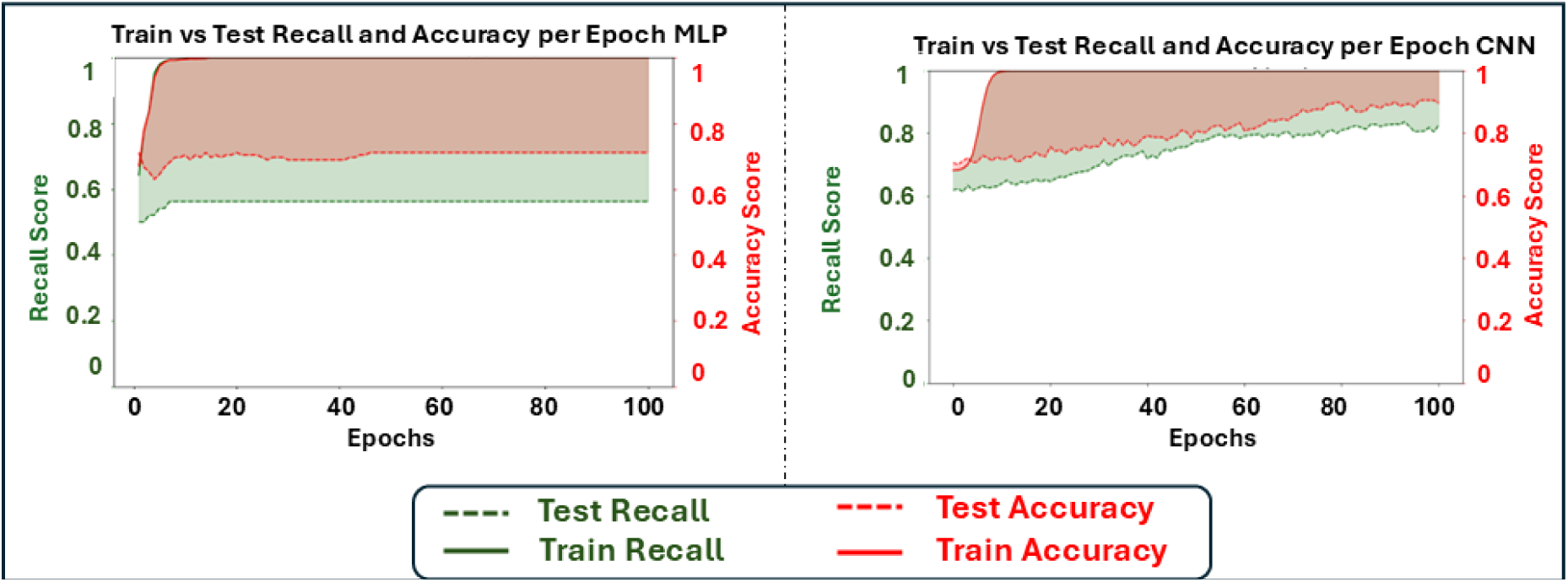
Performance evolution of MLPNN and CNN models following dataset augmentation. Comparison of recall and accuracy metrics across training epochs, with MLPNN results shown on the left and CNN on the right, illustrating the enhanced stability and effectiveness of both models after dataset expansion.

Further analysis was provided by the classification reports (***Fig. 17 C***), offering a quantitative breakdown of the model performance. The MLPNN model exhibited the lowest overall performance among the three models, achieving a precision of 0.77, recall of 0.79, and an F1-score of 0.78 for control cases. For Alzheimer’s cases, the performance dropped notably, with a precision of 0.60, recall of 0.56, and an F1-score of 0.58, highlighting the challenges posed by class imbalance. In contrast, the CNN model demonstrated improved classification metrics for control cases (precision: 0.78, recall: 0.97, F1-score: 0.86), and a moderately better performance for Alzheimer’s cases (precision: 0.89, recall: 0.60, F1-score: 0.64). Although the RF model delivered relatively balanced metrics, control (precision: 0.74, recall: 0.90, F1-score: 0.81), Alzheimer’s (precision: 0.70, recall: 0.52, F1-score: 0.54), its performance remained slightly inferior to the CNN, particularly in Alzheimer’s classification. Overall, SMOTE-based augmentation improved model performance overall, but class imbalance still limited the accurate identification of Alzheimer’s cases.

Finally, the training history plots for the MLPNN and CNN models illustrated their learning dynamics across epochs ***(Fig. 18).*** Both models achieved rapid convergence in training accuracy and recall, reaching values close to 1.0 early in the training process. The MLPNN model demonstrated relatively stable test performance, with consistent recall and accuracy after the initial epochs, indicating effective generalisation. In contrast, the CNN model exhibited higher fluctuations in test recall, though its test accuracy remained comparatively stable across epochs. These results suggest that while both models benefited from synthetic data augmentation in terms of improved training performance, the MLPNN model showed greater stability during evaluation, whereas the CNN’s performance, though strong in accuracy, remained more sensitive to training dynamics.

## Discussion

The primary objective of this study was to develop a classification model capable of distinguishing individuals with AD from healthy controls based on gut microbiome data. As a preliminary step, the gut microbiome was characterised in both groups by analysing diversity metrics and taxonomic composition to identify potential microbial alterations associated with the disease. These insights informed the development of the classification model, aiming to enhance diagnostic accuracy through microbial profiling.

Regarding alpha diversity, our analysis revealed no statistically significant differences in species richness or diversity between groups, as indicated by the Chao1, Shannon, and Simpson indices.

These results suggest that overall microbial diversity and community evenness remain comparable between individuals with Alzheimer’s and controls. Consequently, while microbial composition may differ, there is no clear evidence of a loss of community stability based on alpha diversity metrics. These findings are in line with the original studies from which our dataset was obtained, which also reported no significant differences in alpha diversity. However, previous studies have reported contrasting findings. For example, Zhuang et al. found significantly reduced alpha-diversity in AD patients, with lower Shannon, Simpson, Chao1 indices compared to controls (28). These results suggest that reduced microbial richness and evenness may characterise AD-associated dysbiosis in certain populations. Unfortunately, we were unable to independently assess the findings of Zhuang et al., as the raw sequencing data (e.g., FASTQC files) and associated metadata are not publicly available. Nonetheless, methodological differences, including sequencing platform, preprocessing pipelines, cohort characteristics (e.g., age, diet, geography), and disease stage, could potentially explain the discrepancies observed between studies.

In terms of beta diversity, overall differences in microbial community composition between groups were assessed using Bray-Curtis, Jaccard, and UniFrac distance metrics. Although visual inspection of MDS plots did not show clear separation between AD and control, statistical testing via PERMANOVA revealed significant differences across all distance metrics. The Bray-Curtis index and the Jaccard index indicated small but statistically detectable shifts in microbial composition. Notably, the UniFrac metric, which incorporates phylogenetic relationships, yielded the strongest signal, suggesting that the microbial differences between AD and controls may be more evident at the phylogenetic level than through species presence or abundance alone. These results imply that although group-specific clustering is not visually apparent, AD is associated with subtle compositional shifts in gut microbial communities.

Our findings align in part with those of the referenced studies. In the 16S rRNA-based study (26), no statistically significant differences were detected in beta diversity metrics, nor was there clear visual separation between AD and control groups. In contrast, the WGS-based study (27), did identify significant differences in gut microbial composition between AD and control groups. Using UniFrac-based principal coordinates analysis (PCoA), the WGS-based study detected global taxonomic differences by preclinical AD status. Further analyses, including canonical analysis of principal coordinates (CAP), reinforced these findings, demonstrating that preclinical AD status explained a significant proportion of microbial variation. Notably, these differences were detected at the phylogenetic level, as indicated by the stronger signal observed with UniFrac metrics, while composition-based indices such as Bray-Curtis and Jaccard showed subtler shifts.

Overall, while the referenced study placed greater emphasis on species-level taxonomic associations and functional pathway differences, our results corroborate its broader conclusion that AD is associated with significant yet subtle gut microbiome alterations, detectable through statistical testing but not necessarily evident through visual ordination alone.

At the taxonomic level, the relative abundance of the most common phyla revealed significant differences in *Firmicutes*, *Bacteroidota*, and *Proteobacteria* between AD patients and healthy controls. Notably AD individuals showed higher levels of *Firmicutes* and lower levels of *Bacteroidota*, resulting in an increased *Firmicutes*/*Bacteroidota* ratio. This ratio is commonly considered a marker of gut microbial imbalance, with elevated values often associated with pro-inflammatory states and metabolic dysregulation. In AD, this imbalance may contribute to neuroinflammatory processes and altered gut–brain axis communication. This interpretation is consistent with previous studies in both human and mouse models of AD, which suggest a potential link between gut microbiota imbalance and neurodegeneration (29). Additionally, a significant increase in *Proteobacteria* was observed in the AD group. This phylum is associated with gut dysbiosis and inflammation, and its elevation may reflect gut barrier dysfunction and systemic inflammation, both relevant to AD progression (30). These phylum-level differences were consistent regardless of sex, suggesting that the observed taxonomic changes are likely related to disease status rather than sex-related variation.

Beyond these broad phylum-level differences, genus-level analysis revealed microbial alterations associated with AD. A significant increase in *Dorea* was observed in AD patients compared to controls. Although research on *Dorea* is still limited, preliminary studies suggest that its increased abundance may be linked to cognitive decline. Specifically, higher levels of *Dorea* have been correlated with decreased Mini-Mental State Examination (MMSE) scores, indicating greater cognitive impairment (31). In addition, *Dorea* has been associated with pro-inflammatory pathways, suggesting a potential role in exacerbating neuroinflammatory processes in AD (32). Apart from *Dorea*, two other genera, *Anaerostipes* and *Faecalibacterium*, showed trends approaching statistical significance and were therefore considered noteworthy. An increased abundance of *Anaerostipes* was detected in AD patients. While *Anaerostipes* is generally considered a butyrate-producing genus with anti-inflammatory properties, its specific role in AD remains underexplored (33). Further research is necessary to elucidate its potential involvement in AD-related neuroinflammation. *Faecalibacterium* did not exhibit significant differences between AD and control groups in our study. This observation is important to interpret carefully, as our analysis was conducted at the genus level. This is noteworthy, as *Faecalibacterium prausnitzii*, a key species within this genus, has been associated with cognitive function in prior research. Some studies have reported a decrease in *F. prausnitzii* correlating with cognitive impairment, while others have observed strain-specific effects on cognition. The discrepancy between our findings and those in the literature may reflect the difference in taxonomic resolution, therefore genus-level analyses may mask species-specific effects. Notably, isolated strains of *F. prausnitzii* have been shown to improve amyloid-beta-induced cognitive impairment in mice, suggesting a potential therapeutic role (34). The variability across studies may also be due to differences in study populations, methodologies, or disease progression stages (34). Particular attention was given to the genus *Agathobacter*, a known producer of SCFAs, particularly butyric acid, which has been reported to be reduced in AD patients. Previous research has shown that the abundance of *Agathobacter* negatively correlates with cognitive impairment. Its metabolic activity, particularly through the production of butyrate, appears to have an anti-inflammatory effect that may help counteract neuroinflammatory processes associated with AD. Behavioral studies have further demonstrated that *Agathobacter* supplementation can improve performance in cognitive tasks, suggesting a potential therapeutic benefit (35). In our data, a trend toward increased abundance of *Agathobacter* in male controls compared to females with AD was observed, although this did not reach statistical significance. While most genus-level trends were consistent across sexes, isolated observations like that of *Agathobacter* highlight the need to consider potential sex-specific microbial patterns in future studies.

The second approach used to explore differentially abundant taxa in AD, the LEfSe analysis, further supported our observed taxonomic shifts. It confirmed the enrichment of *Dorea* in the AD group. Additionally, *Faecalibacterium*, which had not reached statistical significance in our initial analysis, was found to be significantly more abundant in AD patients, underscoring its potential relevance in the context of the disease. In addition, this analysis identified a significant reduction in *Ruminococcus* in AD patients compared to controls, a finding consistent with prior research indicating its role in maintaining gut barrier integrity and its potential protective function against neuroinflammation and cognitive decline (36). Similarly, *Clostridium* and *Desulfovibrio* were identified as less characteristic of the AD group, thereby reinforcing prior evidence underscoring their depletion in neurodegenerative conditions (37). Given the established roles of *Clostridium* in gut homeostasis and *Desulfovibrio* in sulphate metabolism, their reduction may contribute to gut dysbiosis and inflammatory processes observed in AD.

Taken together, our findings indicate that alpha diversity does not differ significantly between AD and control groups, suggesting that overall microbial richness and diversity remain comparable, though alternative analytical approaches may be needed to fully explore potential differences. However, beta diversity analyses reveal subtle yet statistically significant shifts in microbial composition, particularly at the phylogenetic level. These ecological changes are reflected in taxonomic alterations, with increased abundance of pro-inflammatory and potentially dysbiotic genera in AD. Altogether, these results point to a biologically meaningful restructuring of the gut microbiome in AD, which may play a role in disease-related inflammatory and neurodegenerative processes.

In parallel with microbiome characterisation, the performance of three distinct machine learning architectures (MLPNN, CNN, and RF) was evaluated for classifying AD and control samples based on microbiome data. Initial model training without data augmentation revealed consistent difficulties in the accurate classification of AD cases across all models, as evidenced by persistently high false negative rates. This trend highlights the inherent complexity of AD-related microbiome signatures and the challenges of learning meaningful patterns from limited, high-dimensional biological datasets.

Among the initially developed models, the RF classifier demonstrated the most robust overall performance, achieving the highest precision for AD samples and both the highest precision and recall for control cases. However, in the context of a diagnostic tool, recall, particularly for AD cases, is crucial, as false negatives (missed diagnoses) might be more harmful than false positives. Nevertheless, recall for AD cases remained low across all models, pointing to a common limitation in the accurate identification of the minority class. The CNN showed competitive performance, especially in terms of precision for AD cases, but fell short in recall. The MLPNN exhibited the weakest performance overall, a shortcoming attributed to overfitting, evidenced by the divergence between training and test recall, despite the application of dropout and regularisation techniques.

To address the limitations of dataset size and class imbalance, synthetic data augmentation using the SMOTE Technique was implemented. This strategy led to overall improvements in classification performance, particularly enhancing recall for the underrepresented Alzheimer’s class. Among the models, CNN and RF showed the best results, while MLPNN, although less performant, also benefited from the augmentation. These findings highlight the effectiveness of SMOTE in mitigating class imbalance and improving the models’ ability to detect Alzheimer’s cases.

However, it is important to acknowledge that, while SMOTE improved performance metrics, it does so by generating synthetic instances that replicate existing data distributions. In essence, it simulates rather than introduces genuinely novel data. Consequently, although SMOTE offers valuable insights into how the models might behave with a larger dataset, it does not fully replicate the variability and complexity that real-world data would provide. This observation reinforces the notion that data augmentation can serve as a useful proxy for assessing model scalability, but it cannot substitute the value of actual, diverse samples.

Given this context, we argue that the observed improvements, particularly in recall, support the hypothesis that larger, real-world datasets would lead to substantial performance gains. Notably, the CNN’s behaviour under synthetic augmentation suggests a stronger potential for generalisation than the RF or MLPNN models. While the RF classifier may be preferable in initial scenarios due to its lower computational cost and solid performance on limited data, we support the hypothesis that, with access to more extensive and diverse real-world datasets, the CNN would ultimately surpass RF in diagnostic accuracy. This is especially relevant in early diagnosis scenarios, where the ability to generalise from complex patterns is essential.

### Limitations

Several limitations should be acknowledged in this study. Firstly, the relatively small sample size limits the generalisability of the findings and reduces the statistical power to detect subtle microbiome differences associated with AD. Moreover, the limited amount of data constrains the full potential of AI models, particularly neural networks, which typically require large, diverse datasets to achieve optimal performance and avoid overfitting. Secondly, the datasets were compiled from independent studies, each employing different sequencing methodologies (16S or WGS), introducing technical variability and batch effects despite efforts to correct for them. Currently, there is also a lack of standardised protocols to enable the effective integration of 16S and WGS data, which further exacerbates these batch effects and may introduce bias or error into downstream analyses. Thirdly, all samples analysed were derived from faecal material, which, while widely used in microbiome studies, is susceptible to lifestyle related influences such as diet, medication, and geographic factors, adding variability unrelated to disease status. Finally, the inherent complexity of linking gut microbiome alterations to a multifactorial disease like AD must be emphasised, given the many biological, environmental, and genetic factors involved in gut–brain axis interactions. These limitations underscore the necessity for future studies involving larger, more homogeneous cohorts, standardised sequencing protocols, and integrated multi-omics approaches to better elucidate the relationship between the gut microbiome and AD.

## Conclusions

- AD is associated with subtle but significant gut microbiome alterations.
- Although integrating 16S rRNA and WGS data through a unified pipeline using a common taxonomic reference reduced batch effect, a residual batch effect was still observed.
- While alpha diversity metrics indicated no significant differences in richness or evenness between AD and controls, beta diversity analyses revealed compositional shifts, suggesting alterations in microbial community structure in AD.
- AD patients exhibited increased abundance of pro-inflammatory or potentially dysbiotic taxa such as *Dorea*, *Proteobacteria*, and a higher *Firmicutes*/*Bacteroidota* ratio.In contrast, healthy controls showed higher levels of taxa with anti-inflammatory or neuroprotective potential, including *Agathobacter*, *Clostridium*, and *Desulfovibrio*.
- Machine learning models trained on microbiome data were able to distinguish AD from controls with moderate accuracy, with the RF showing the best overall performance.
- SMOTE-based augmentation improved classification metrics, particularly recall, with the CNN showing the best overall performance.

## Competing interests

E. C-SP and L. J. M-Z are co-founders and E.C-SP Data Science Director (part time) of Microsei Biotech S.L.

## Author Contributions

A. P.-C. formal analysis, investigation, methodology, visualization, writing—original draft preparation. B.L.-P. methodology. D. C.-C. methodology. L.J.M.-Z. funding acquisition, methodology, writing—review & editing. E.C. d S.P. funding acquisition, project administration, supervision, writing—review & editing. A. M.-S. conceptualization, methodology, project administration, supervision, writing—original draft preparation.

## Funding

This work has been carried out under the context of the projects AI4FOOD (Y2020/TCS-6654) and TEC-2024/BIO-167 CD3DTech-CM (ORDER 5696/2024, B.O.C.M. N°. 307 12/26/2024 both funded by Community of Madrid. Project PID2023-150146OA-I00 founded by MICIU/AEI/10.13039/501100011033 and FEDER, UE. Institute of Health Carlos III (project IMPaCT-Data, exp. IMP/00019), co-funded by the European Union, European Regional Development Fund (A way to make Europe’); RED2022-134934-T funded by MICIU/AEI/10.13039/501100011033. COST Actions CA18131 - ML4Microbiome, CA23110 - INFOGUT and CA20128 - PIMENTO. A.P-C. is funded by Programa FSE+2021-2027 (AI2025/002-PEJ-2024-AI/COM-32727). B.L.-P is funded by Formación del Profesorado Universitario grant (FPU22/04053) funded by MICIU/AEI/10.13039/501100011033. D.C-C. is funded by “Programa de Jóvenes Investigadores” (09-PIN1-00014.8/2024). A. M.-S is funded by the European Union (MSCA, Ref.: 101105645).

## Data Availability

All data used in the present study was aready publicly available at ENA Browser (https://www.ebi.ac.uk/ena/browser/home).

https://github.com/albaperezcuervo/TFM/tree/main

https://www.ebi.ac.uk/ena/browser/view/PRJNA798058

https://www.ebi.ac.uk/ena/browser/view/PRJNA801673

## Notes

### Author Declarations

ENA Browser (https://www.ebi.ac.uk/ena/browser/home)

